# Epigenetic patient stratification reveals a sub-endotype of type 2 asthma with altered B-cell response

**DOI:** 10.1101/2025.08.28.25334696

**Authors:** Aditya Gorla, Jonathan Witonsky, Zeyuan Johnson Chen, Jennifer R. Elhawary, Joel Mefford, Javier Perez-Garcia, Anne-Marie Madore, Scott Huntsman, Donglei Hu, Celeste Eng, Nirav R. Bhakta, Prescott G. Woodruff, Catherine Laprise, Sriram Sankararaman, Jonathan Flint, Christopher D. C. Allen, Elad Ziv, Noah Zaitlen, Esteban Burchard, Elior Rahmani

## Abstract

Despite biomarker-guided treatment strategies, clinical outcomes among patients with type 2 (T2)-high asthma remain heterogeneous, with some patients responding poorly to T2-targeted biologic therapies. We developed a contrastive machine learning method for patient stratification based on whole-blood DNA methylation (DNAm), applying it to pediatric asthma cohorts of Latino (discovery; n=1,016) and African American (replication; n=429) children. The resulting DNAm stratification score revealed a continuum of clinical severity and drug response within the T2-high asthma endotype. Molecular profiling of high-score asthma patients identified eosinophil-specific hypermethylation—validated in an independent Canadian adult cohort using purified eosinophil DNAm—as well as upregulation of canonical T2-associated genes. Transcriptomic analysis of elevated DNAm scores within T2-high patients further uncovered a gene signature linked to B-cell lineage activity, predominantly reflecting plasma cell activity orthogonal to canonical T2 inflammation programs. This defines a previously unrecognized sub-endotype, which we term T2-high asthma with Altered B Cell response (T2ABC). In a randomized controlled trial of the anti-IgE biologic omalizumab in primarily White adult T2-high asthmatic patients (n=300), the T2ABC gene expression signature was prognostic of poor outcomes, including a 24% mean increase in disease exacerbation rates compared to the trial baseline (P=0.004), which could not be explained by treatment or placebo assignment. Patients treated with omalizumab showed better outcomes than patients in the placebo arm within the T2ABC-low group (P=0.019) but not within the T2ABC-high group (P=0.48), suggesting that IgE blockade does not adequately target the pathogenic mechanisms active in T2ABC-high disease. Single-cell transcriptomic analysis demonstrated that the T2ABC signature reflects heightened activity of non-IgE plasma cells, consistent with the presence of additional antibody isotype responses in a form of severe asthma arising within a T2-high immunologic context. Our findings, replicated and validated across four ancestrally and ethnically diverse pediatric and adult cohorts, support the use of DNAm- and transcriptome-based patient stratification to refine drug development, eligibility, and administration strategies for improving precision in T2 asthma therapy.

## Introduction

Asthma is a globally prevalent, high-burden disease marked by substantial clinical heterogeneity and variable treatment response^1,2^. This heterogeneity arises from interindividual differences in disease mechanisms and their clinical presentations, shaped by genetic predisposition^3^, environmental exposures^4^, and their interactions^5^. Defining biologically and clinically meaningful subtypes is central to precision medicine efforts in asthma. Molecularly defined subtypes—known as endotypes—have been associated with disease severity and clinical trajectory^6^. Although endotype-based classification has advanced asthma care, treatment responses remain inconsistent, and disease control suboptimal for many patients^7,8^.

Among these endotypes, type 2 (T2)-high and T2-low asthma are distinguished by underlying immune mechanisms and inflammatory profiles, each associated with biomarkers that guide therapy^9^. T2-high asthma is primarily driven by type 2 helper T (Th2) cells and group 2 innate lymphoid cells (ILC2s), which secrete interleukins (IL)-4, IL-5, and IL-13^9,10^. These cytokines orchestrate allergic and eosinophilic inflammation, manifesting clinically as allergic sensitization and elevated blood eosinophil counts (BEC)^9,10^. In contrast, T2-low asthma may in part be characterized by neutrophilic or paucigranulocytic airway inflammation, often with lower levels of allergic sensitization and systemic eosinophilia relative to T2-high asthma^9,10^. While paucigranulocytic asthma typically presents with milder disease, the neutrophilic subtype is strongly associated with severe, treatment-resistant asthma^11^. Biomarker-based endotype classification using BEC, sputum eosinophils, fractional exhaled nitric oxide (FeNO), and total serum immunoglobulin E (IgE) supports personalized use of targeted biologic therapies^10^.

Endotype definitions capture broad immune profiles but often fail to resolve finer-grained variation in treatment response. In particular, even when patients meet biomarker thresholds for T2 inflammation, responses to targeted biologics—often exceeding $40,000 annually—are frequently incomplete^12,13^. In a large clinical study, fewer than 25% of adults with severe asthma achieved clinical remission despite biologic therapy^14^. Similarly, among adults with elevated IgE and allergic sensitization, 20-30% do not respond to treatment with omalizumab, a monoclonal antibody that targets IgE^15,16^. These findings highlight the limitations of current biomarkers for predicting treatment response and underscore the need for more precise stratification tools to uncover previously obscured disease mechanisms and inform novel therapeutic approaches.

Both recently identified and yet-to-be-recognized molecular heterogeneity within established asthma endotypes undermines the precision of existing biomarker-guided therapy^17^. Emerging data indicate that individuals classified within the same T2-high endotype may differ markedly in immune programs, cellular mediators, and treatment response^18^. This heterogeneity extends beyond biologics: African American and Puerto Rican children—two populations with disproportionately high asthma prevalence and morbidity—exhibit reduced bronchodilator response (BDR) to short-acting β2-agonists (SABAs)^19^, even when combined with inhaled corticosteroids (ICS)^20^. Clinical heterogeneity, compounded by population-level variation in biomarker distributions, further compromises asthma care and perpetuates inequities through biased endotype classification thresholds and treatment eligibility criteria^21,22^.

Advancing both precision and equity in asthma care requires clinically practical, cost-effective, and unbiased tools that can stratify patients beyond existing biomarker frameworks. Achieving this goal necessitates computational approaches that can uncover *de novo*, clinically meaningful patient subgroups from high-dimensional data. In particular, genetic, epigenomic, and transcriptomic datasets offer powerful means to uncover latent disease structure and capture immune heterogeneity not reflected in conventional biomarkers^3,23–25^. Yet, most existing computational methods rely on clustering or dimensionality reduction, assuming these methods will reveal explanatory patterns corresponding to disease subtypes. While this has shown value in select cases^26–29^, it faces a fundamental limitation that restricts its generalizability. These techniques essentially group patients by overall similarity, treating all features as equally informative. As a result, disease subtypes characterized by subtle molecular signals are likely to be obscured by dominant sources of variation—patterns that are correlated with many features. In genomic data, such dominant variation often reflects confounding factors like cell-type composition, batch effects, ancestry, or unknown ascertainment or experimental biases, rather than true disease heterogeneity.

To overcome the limitations of existing methods, we introduce a novel contrastive machine learning method for patient stratification. We apply this method to whole-blood DNA methylation (DNAm)—a set of epigenetic modifications that influence immune cell function, drive allergic inflammation, and are associated with asthma pathogenesis and treatment response^30^—to define a patient stratification score for asthma. This DNAm score reveals a clinically meaningful continuum of disease severity and therapeutic response within the T2-high asthma endotype. Importantly, it identifies a distinct molecular sub-endotype—T2-high asthma with altered B cell response (T2ABC)—characterized by differential expression of genes related to B cells and plasma cells within a T2 inflammatory context.

## Results

### A novel contrastive machine learning approach for uncovering disease heterogeneity

We developed Phenotype Aware Component Analysis (PACA), a novel contrastive machine-learning method for defining biomedically meaningful disease subtypes and patient stratifications from high-dimensional data. Unlike standard unsupervised approaches (e.g. principal components analysis), PACA does not simply cluster patients based on the dominant variation in the data. Instead, PACA isolates disease-specific heterogeneity by first removing sources of variation that are shared with healthy control individuals from the disease case data, and then applies standard dimensionality reduction techniques to the resulting disease-specific signal. By accounting for sources of variation present in both cases and controls, we expect the top axes of variation in case data to reflect disease heterogeneity (Fig. 1a). Since these shared sources of variation are generally unknown, we take an unsupervised approach to identify latent features that are consistent across the two groups. Unlike existing contrastive learning methods^31,32^, PACA does not require tuning a non-intuitive contrastive hyperparameter that controls the level of variation retained among cases relative to controls. See Methods and Supplementary Note S1 for more details. We conducted comprehensive benchmarking and confirmed that PACA is statistically calibrated and better powered to identify disease heterogeneity compared to other contrastive learning methods^31,32^ when applied to synthetic and real-world data, including gene expression, DNAm, and genotype data (Supplementary Note S2; Supplementary Fig. S1-S5).

**Fig. 1:**
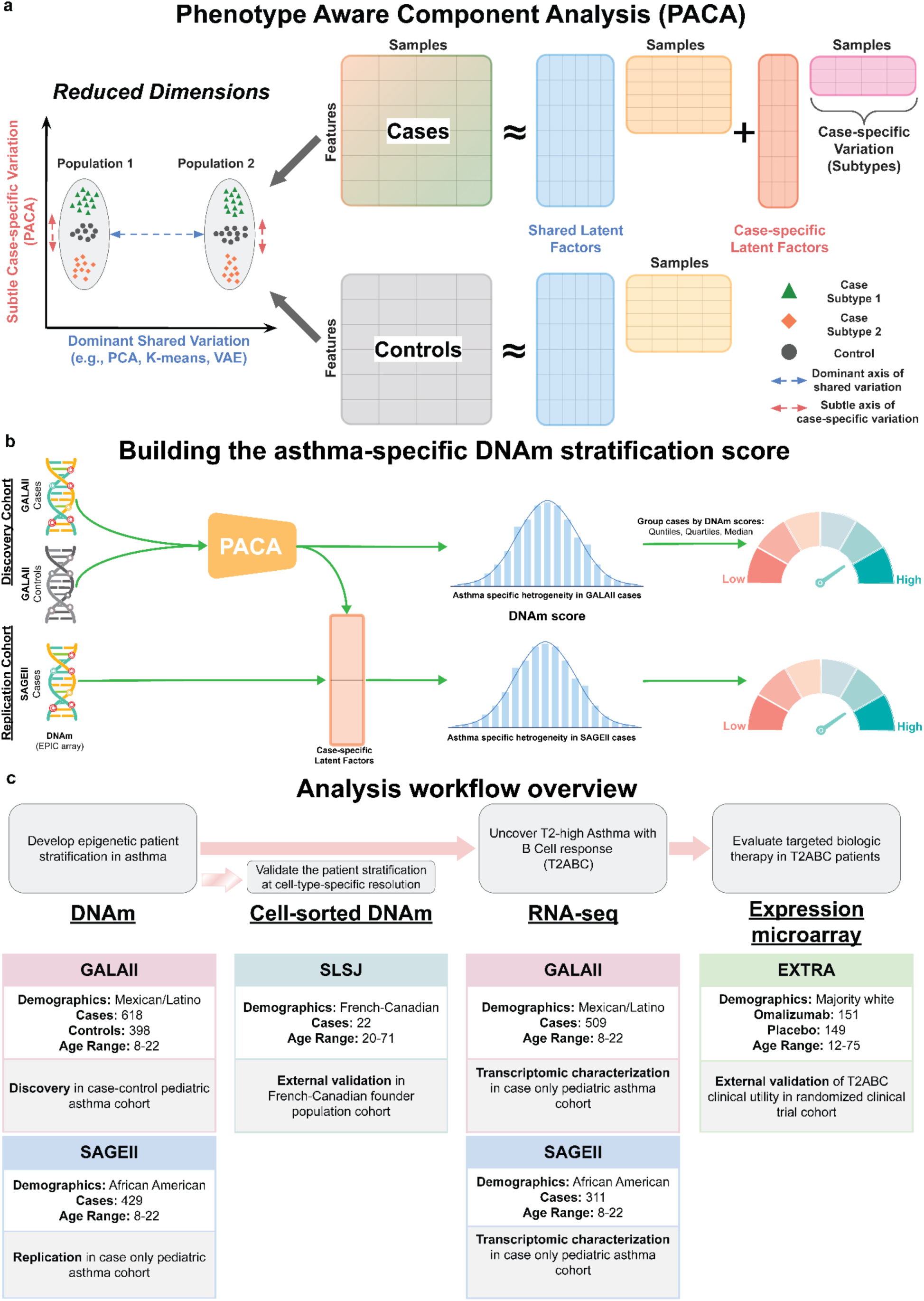
Overview of PACA and the study design for uncovering clinically relevant patient stratification within T2-high asthma. **(a)** PACA models both shared latent factors present in cases and controls and case-specific variation (right). Unlike standard unsupervised dimensionality reduction methods, which typically recover dominant axes of variation, such as population structure or technical artifacts, PACA isolates subtle, case-intrinsic variation that is more likely to reflect disease heterogeneity (left). **(b)** Schematic of PACA applied to asthma case-control DNAm data to derive an asthma-specific DNAm stratification score that reflects heterogeneity among cases. **(c)** Overview of the cohorts used in the multi-modal analysis workflow, including DNAm, RNA-seq, and microarray expression data across diverse populations for discovery, replication, and validation analyses.

### An epigenetic patient stratification score captures clinically significant asthma heterogeneity

We hypothesized that DNAm will be an effective input for robust patient stratification: in contrast to static germline variants, DNAm signatures are dynamic—evolving across the life course and associated with disease states—yet exhibit greater stability than RNA-based measures^33,34^. We utilized PACA to develop an asthma patient stratification score using whole-blood DNAm from Genes-environments & Admixture in Latino Americans II (GALA II), a pediatric asthma cohort primarily of Mexican and Puerto Rican ethnicities (n=618 cases, n=398 controls; Supplementary Note S3; Fig. 1b,c and Supplementary Table S1). To reduce noise and narrow the feature space in the data, we first evaluated all methylation sites passing quality control (751,876 cytosine-phosphate-guanine probes, or CpGs) for statistical association with asthma (Methods). An analysis with PACA considering only CpGs that demonstrated a nominally significant association with asthma (P<0.01) resulted in a patient stratification model based on a linear combination of 7,662 CpGs (Supplementary Table S2). To confirm the cross-population consistency of the resulting asthma stratification, we applied the 7,662-CpG DNAm score to a pediatric African American patient replication cohort with whole-blood DNAm from the Study of African Americans, Asthma, Genes, and Environments (SAGE II; n=429 cases; Fig. 1b,c and Supplementary Table S1).

In both GALA II and SAGE II, the DNAm score stratified patients along a continuum corresponding to heterogeneity in lung function (spirometry) and clinical presentation of asthma (Fig. 2a,b). Specifically, high DNAm scores are associated (Pearson correlation; Bonferroni adjusted P<0.05) with lower baseline lung function as measured by forced expiratory volume in one second (FEV1), forced vital capacity (FVC), peak expiratory flow rate (PEF), and forced expiratory flow (FEF). Other asthma phenotypes in correlation with high DNAm scores include higher BDR, elevated BEC and IgE, and higher exacerbation scores (Methods; Fig. 2a,b and Supplementary Tables S3 and S4).

**Fig. 2:**
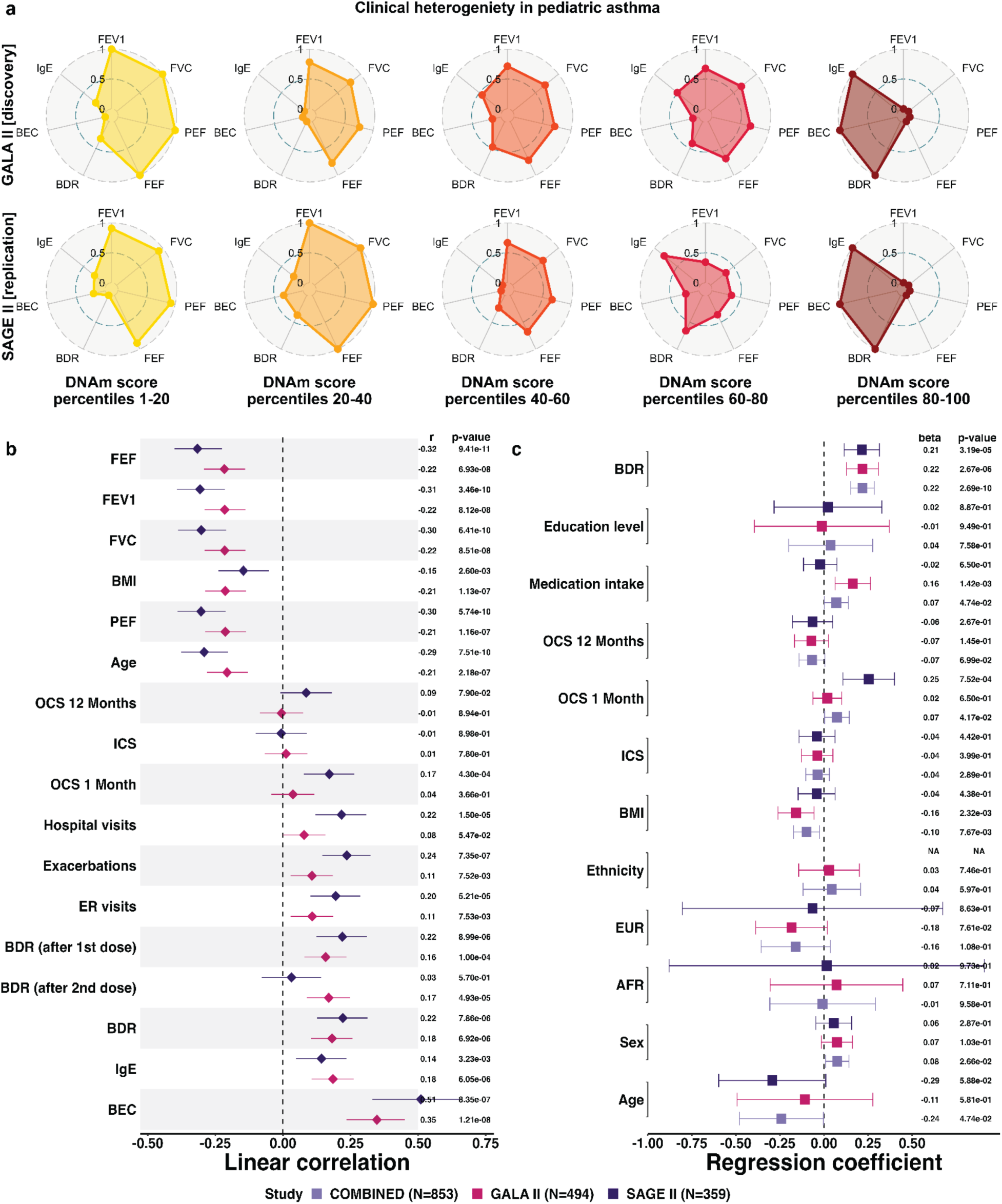
PACA DNAm scores stratify patients along a continuum corresponding to heterogeneity in the clinical presentation of asthma. **(a)** Radar plots illustrating distribution shifts of asthma phenotypes across different percentile ranges of the DNAm score. Each point on the radial scale represents a mean phenotypic value for the respective quantile range, normalized to a 0 to 1 scale within each cohort. **(b)** Pearson’s coefficients and 95% confidence intervals (CIs) for the correlation between the DNAm scores and clinical covariates. **(c)** Linear regression coefficients and 95% CIs for the DNAm score as the outcome and BDR as the variable of interest, adjusted for demographic and clinical variables. Results are stratified by study (GALA II, SAGE II) or combined (COMBINED). Abbreviations: AFR: African ancestry (%), BDR: bronchodilator response (%), BEC: blood eosinophil count (cells/μL), BMI: body mass index, ER: emergency room; EUR: European ancestry (%), FEF: forced expiratory flow (% predicted), FEV1: forced expiratory volume in 1 second (% predicted), FVC: forced vital capacity (% predicted), ICS, inhaled corticosteroids, IgE: serum total immunoglobulin E (log-transformed), OCS: oral corticosteroids, PEF: peak expiratory flow (% predicted).

The associations with BDR, BEC, and IgE remained significant (linear regression; GALA II P<2.7e-6, SAGE II P<2.9e-3) after adjusting for demographics (age, sex, ancestry, and ethnicity), body mass index (BMI), education level, medication intake, and inhaled (ICS) and oral (OCS) corticosteroids use (Fig. 2c and Supplementary Fig. S6). Inspecting the top 100 most informative CpGs of our stratification model revealed heavily weighted CpGs in genes implicated in asthma pathogenesis and regulation of airway inflammation and remodeling (*STAT3*, *RASSF1*, *MEOX1*)^35–37^, bronchodilator response (*DDX54*)^38^, and asthma severity and lung function (*ALDH2*)^39,40^ (Supplementary Table S2). We confirmed that existing contrastive learning methods did not yield similarly meaningful asthma patient stratification (Supplementary Note S2.6; Supplementary Tables S3 and S5).

### BEC and IgE predict bronchodilator response only in patients with high DNAm scores

We evaluated whether our patient stratification model explains variability in biomarker-based prediction of clinical outcomes, first focusing on BDR to the SABA albuterol, the most used drug for acute bronchospasm worldwide. The widely used asthma biomarkers BEC and IgE, both found to be associated with our DNAm score (Fig. 2a,b), are known to be predictive of BDR^41^. Across GALA II and SAGE II, the odds ratio (OR) for drug response (BDR≥12%)^42^ is 1.14 (95% CI [1.10, 1.18]) and 1.23 (95% CI [1.12, 1.35]) for BEC and IgE, respectively (Supplementary Fig. S7).

We investigated if the DNAm score stratifies the population such that these biomarkers are predictive of BDR only in specific patient subgroups along the stratification spectrum (i.e., a statistical interaction). A linear regression model for BDR as the outcome revealed a significant interaction between IgE levels and the DNAm score (GALA II P=0.005, SAGE II P=3.3e-6) after adjusting for IgE levels, demographics, BMI, education level, medication intake, ICS, and OCS (Supplementary Fig. S8). Similarly, we found a significant interaction between BEC and the DNAm score in GALA II (P=5.2e-5; Supplementary Fig. S8). This result was not replicated in SAGE II, likely due to the limited number of BEC measurements available in this cohort (n=83). Even so, addressing the sample size limitation by imputing missing BEC proportions (i.e., eosinophils as a percentage of leukocytes) from DNAm levels^43^ (Methods; Supplementary Fig. S9) revealed a significant interaction between BEC and our DNAm score, which could not be explained by the BEC proportions themselves or other covariates (GALA II P=0.008, SAGE II P=4.4e-4; Supplementary Fig. S8). Hereafter, we consider BEC proportions rather than counts, unless indicated otherwise.

The statistical interactions suggest that elevated BEC and IgE predict BDR only in patients with high DNAm scores. Indeed, in GALA II, classifying patients into responders (BDR≥12%) and non-responders, IgE levels correlate with drug response in patients with above-median methylation scores (OR for response 1.42, 95% CI [1.24, 1.63]; P=3.9e-7) but not in patients with below-median scores (OR 1.05, 95% CI [0.92, 1.2]; P=0.57); BEC correlates with response in upper-quartile (OR 1.12, 95% CI [1.04, 1.22]; P=7.9e-4) but not in lower-quartile patients (OR 1.05, 95% CI [0.95, 1.17]; P=0.21) (Fig. 3a). More generally, we observe a linear increase in the correlation of BEC and IgE with drug response along the DNAm score continuum, which replicates in SAGE II (Fig. 3a,b and Supplementary Fig. S10). Since low baseline lung function may explain BDR^41^, we further verified that our results hold when restricting the analysis to this subgroup (Methods; Supplementary Fig. S11). Overall, our results confirm a robust monotonous increase in the effectiveness of elevated BEC and IgE as predictive drug response biomarkers along the patient stratification spectrum.

**Fig. 3:**
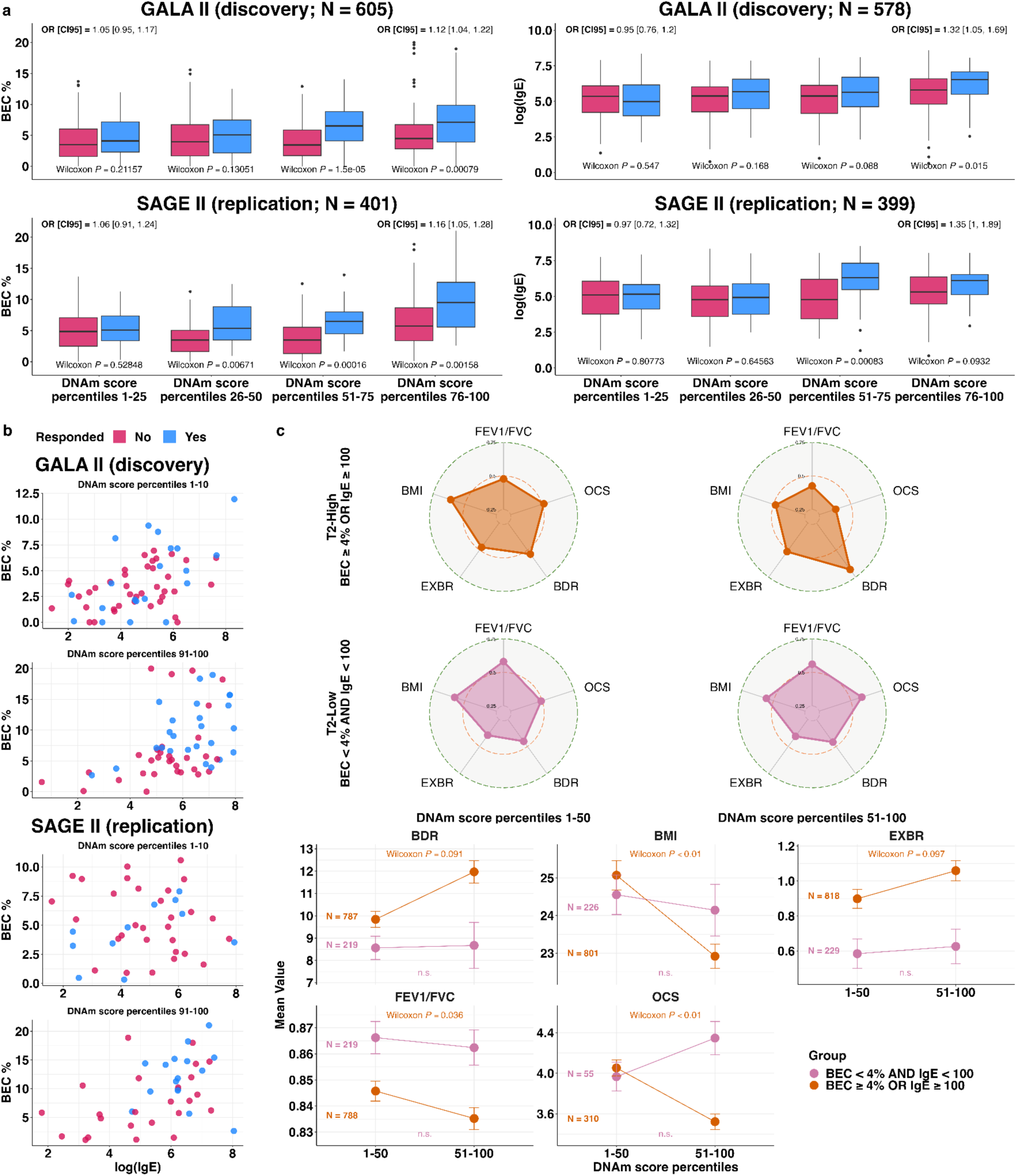
BEC and IgE as drug response (BDR) biomarkers along the DNAm patient stratification spectrum. **(a)** Grouped boxplots comparing the distributions of imputed blood eosinophil proportions (BEC%; left) and (log) IgE levels (right) across different percentile ranges of the DNAm score, stratified by BDR status. A low Wilcoxon rank-sum p-value and a high odds ratio (with 95% confidence intervals) indicate the biomarker is predictive of BDR for patients in the respective percentile range. **(b)** The empirical joint distribution of BEC% and (log) IgE in patients with top and bottom decile DNAm scores, stratified by BDR status. **(c)** Distribution shifts in clinical phenotypes associated with asthma severity. Top: radar plots comparing T2-high (orange) and T2-low (purple) patients, stratified by below- and above-median DNAm scores. Each point on the radial scale represents a mean phenotypic value, normalized to a 0 to 1 scale within each plot. Data are pooled across GALA II and SAGE II to increase statistical power. Bottom: mean ± standard error for each clinical phenotype in the radar plots, evaluated separately for the four patient subgroups. P-values are based on Wilcoxon rank-sum tests comparing patients with below- and above-median DNAm scores. A significant p-value in only one of the two endotypes (T2-high or T2-low) suggests a statistical interaction. Abbreviations: EXBR: Exacerbation score in the last 12 months (0-6); OCS: Time since the last oral steroid prescription (a lower value indicates a more recent prescription).

Next, we evaluated the performance of logistic regression models using both BEC and IgE as two-biomarker models for BDR prediction (Methods; Supplementary Table S6). A standard two-biomarker model based on the entire patient population yielded receiver operating characteristic area under the curve (ROC AUC) of 0.62 and 0.69 on GALA II (train) and SAGE II (test), respectively. In contrast, a two-biomarker model for patients with top-decile DNAm scores achieved ROC AUC of 0.76 and 0.75 in GALA II and SAGE II, respectively. As expected, a two-biomarker model for patients with bottom-decile DNAm scores, for which BEC and IgE are not associated with BDR, performed poorly (ROC AUC of 0.62 and 0.59). Evaluating other percentile ranges demonstrated a linear increase in performance along the DNAm score continuum (Supplementary Table S6). Alternative predictive models for BDR, including a model based on cell-type composition and a model based on the same CpGs used in our DNAm score, underperformed compared to a two-biomarker model tailored for patients with high DNAm scores (Supplementary Note S4; Supplementary Table S6).

### The DNAm score reflects heterogeneity of T2 asthma characterized by differential eosinophil-specific methylation

The association of high DNAm scores with T2-high asthma (elevated BEC and IgE; Fig. 2a,b) could suggest that our DNAm score merely recapitulates known T2 endotypes. However, the DNAm score stratified the effectiveness of BEC and IgE in predicting BDR, even when the analysis was restricted to T2-high patients (BEC≥4% or IgE≥100 kU/L), as defined by elevated BEC or IgE levels (Methods; Supplementary Fig. S11), an effect not observed among T2-low patients (BEC<4% and IgE<100 kU/L; Supplementary Fig. S12). Moreover, within the T2-high group, higher DNAm scores were associated with established clinical markers of asthma severity—including increased exacerbation frequency, BMI, more recent OCS use, and reduced FEV1/FVC ratio^44^—associations that were not observed among T2-low patients (Fig. 3c). Together, these findings suggest that the DNAm score primarily represents unknown variation that pertains to T2 asthma.

To better understand the biological basis of this variation within T2 asthma, we next sought to profile the molecular features associated with the DNAm-based patient stratification, with particular attention to cell-type specificity. Using TCA, a deconvolution method for bulk DNAm^45^, we tested our model’s top 100 most informative CpGs for cell-type-level differential DNAm with the DNAm scores, while accounting for demographics, technical variation, and cell-type composition (Methods). We identified nine CpGs associated with the DNAm score, all of which are specific to eosinophil cells (Supplementary Table S7). These CpGs, identified in GALA II (FDR<0.05) and replicated in SAGE II (P<0.05), mark hypermethylation in eosinophils of samples with high DNAm scores. Eight of these nine eosinophil-specific associations were identified and replicated within the T2-high patient group, but none were observed in T2-low patients (Supplementary Tables S8 and S9). Two of the replicating CpGs, cg18254848 and cg26724455, have previously been associated with allergic airway inflammation and childhood asthma: cg18254848, located in the *CLC* gene, has been associated in whole-blood DNAm data with IgE levels, allergic asthma, and increased FeNO^46^; cg26724455, located in *VTI1A*, has been reported as differentially methylated in nasal epithelial cells from children with asthma and high FeNO^47^.

Our results suggest that eosinophil-specific epigenetic programming may underlie the variation within T2 asthma captured by our DNAm stratification model. However, these findings are based on computational deconvolution of whole-blood DNAm data, which has known limitations^45,48^. To independently validate our results, we analyzed DNAm profiles from isolated eosinophils collected from 24 individuals (16 cases and 8 controls) in the Saguenay–Lac-Saint-Jean (SLSJ) asthma family cohort, comprising primarily adults from a French-Canadian founder population with available spirometry^49,50^. For each sample, we computed a composite hypermethylation score by summing methylation levels across the available eosinophil-specific CpGs identified in our primary analysis (Methods). We first confirmed that the eosinophil composite scores were associated with asthma case-control status (P=0.014). Second, although the small sample size presumably limited our ability to detect associations with several lung function metrics, we observed a significant association with FEV1/FVC (Bonferroni-adjusted P<0.05; Supplementary Table S10), consistent with our primary analysis (Fig. 3c). Notably, this association was identified despite the absence of sufficient BEC and IgE data to define T2-high status in this cohort, which further reduced statistical power. These findings support the interpretation that our DNAm stratification captures clinically meaningful variation through eosinophil-specific hypermethylation.

### Elevated DNAm score marks a sub-endotype within T2-high asthma characterized by an altered B cell response

The DNAm score is associated with clinical and molecular heterogeneity within T2-high asthma; however, the methylation differences the score reflects may not represent direct causal mechanisms. Rather, it may statistically tag upstream transcriptional programs or regulatory variation that drive functional differences. This prompted us to further investigate how variation in the DNAm score corresponds to gene expression changes. Using whole-blood RNA sequencing (RNA-seq) data collected from the same GALA II and SAGE II participants, we first performed a genome-wide differential gene expression analysis to identify genes associated with the DNAm score (Methods). We identified 196 replicating differentially expressed genes (DEGs) across GALA II and SAGE II (FDR < 0.05 in both cohorts independently), adjusting for demographics, education level, ICS, OCS, and technical variation using SVA^51^ (Supplementary Tables S11 and S12). Gene ontology enrichment analysis^52^ indicated a significant overrepresentation of gene sets related to inflammatory response, T cell activation, and regulation of activated T-cell proliferation (FDR<0.05; Supplementary Table S13).

Consistent with the association between high DNAm scores and T2-high asthma, we observed overexpression of canonical T2 genes implicated in the pathways or directly targeted by current biologic therapeutics, including *IL1RL1*, *CCR3*, and *IL5RA*^10,53,54^. However, the DNAm score was associated with distinct clinical and epigenetic variation within the T2-high group, suggesting that it captures heterogeneity beyond the current definition of the T2-high endotype. To dissect this further, we performed a 2-way differential expression analysis, conceptually similar to a method used to uncover genetic heterogeneity within disease subtypes^55^ (Fig. 4a). First, we tested for DEGs between T2-high and T2-low patients (“T2 endotype DEGs”), and second, we tested for DEGs associated with the DNAm score within the T2-high group (“T2-high DEGs”; Methods).

**Fig. 4:**
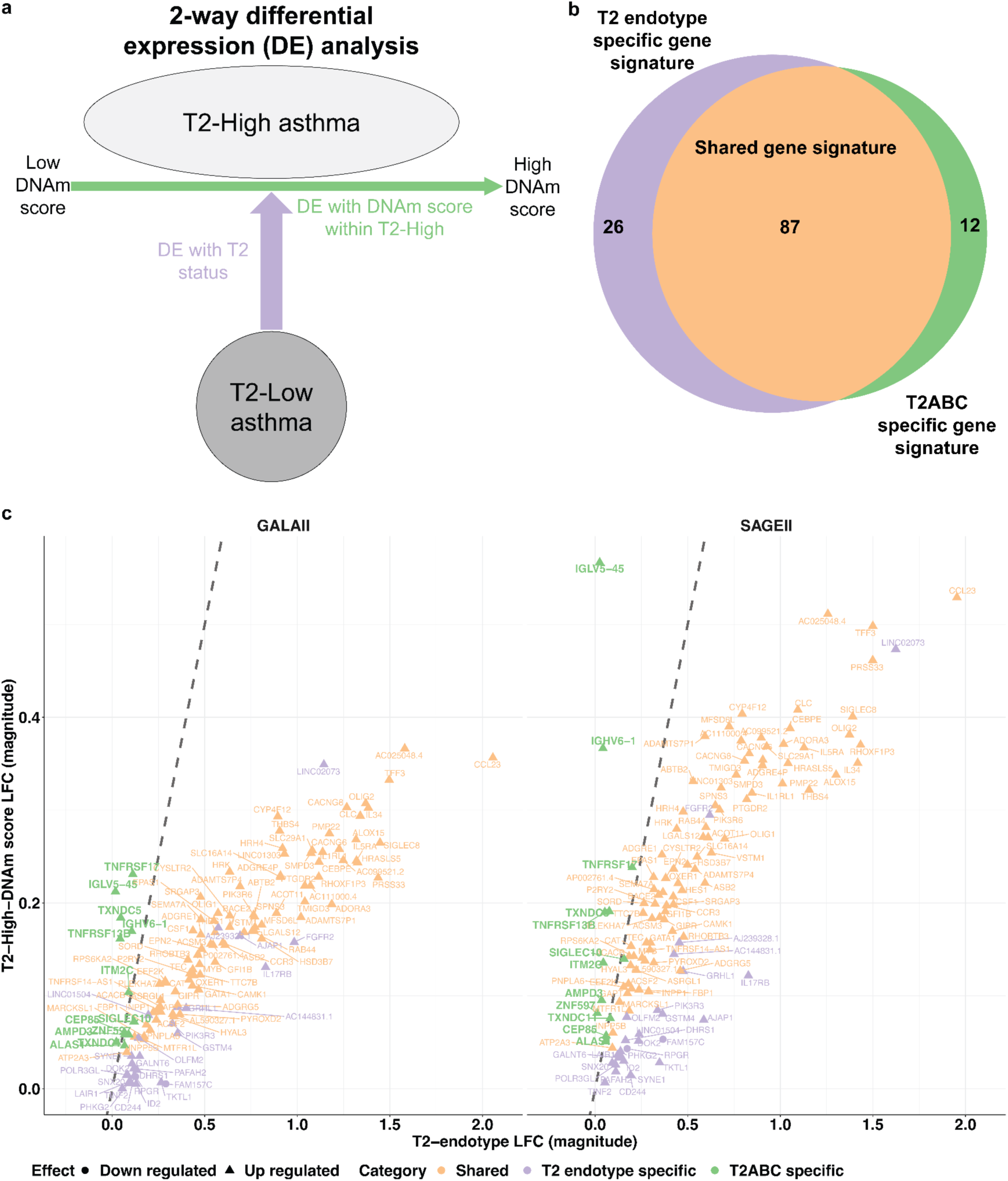
The DNAm-based score reveals a B-cell response sub-endotype within T2-high asthma. **(a)** Schematic of 2-way differential gene expression analysis. Genes were tested for association with T2 status (T2-high versus T2-low) and, separately, for association with the DNAm score within the T2-high group. **(b)** Venn diagram showing the number of DEGs (FDR<0.05 in both GALA II and SAGE II) unique to the T2 status comparison (purple), unique to the DNAm score within T2-high asthma (green; T2ABC signature) or shared between both analyses (orange). **(c)** Scatter plots showing the log fold change (LFC) from the T2 status comparison (x-axis) versus the LFC from the DNAm score association within T2-high asthma (y-axis), in GALA II (left) and SAGE II (right). Genes are color-coded as in (b).

This 2-way analysis resulted in two (overlapping) gene sets (Fig. 4b,c). We found 87 DEGs shared between the two gene sets, 26 genes that are unique to the set of T2 endotype DEGs, and 12 genes that are unique to the set of T2-high DEGs (FDR<0.05 in both GALA II and SAGE II independently; Supplementary Tables S14 and S15). The shared DEGs included canonical T2 markers, such as *IL5RA*, *IL1RL1*, *CYSLTR2*, and *SIGLEC8*. Notably, the 12 T2-high-specific but T2-pathway-independent DEGs were all overexpressed in patients with elevated DNAm scores (Supplementary Tables S14 and S15). Eight of these genes are functionally linked to B-cell lineage populations: *SIGLEC10*, which is involved in B cell regulation and broader immunomodulatory roles across other immune cell types, including T cells and dendritic cells^56–58^; *TNFRSF13B*, a dual-function regulator that constrains excessive B cell proliferation while promoting plasma cell differentiation and survival^59^; and six others—*TNFRSF17*, *IGHV6-1*, *IGLV5-45*, *ITM2C*, *TXNDC5*, and *TXNDC11*. These six genes are predominantly expressed in plasma cells, based on whole-blood deconvolution^60^ of the GALA II and SAGE II datasets (Supplementary Note S5; Supplementary Table S16) and by analysis of eight independent single-cell RNA-seq datasets from human blood and intestine, where plasma cells are naturally more abundant (Methods; Supplementary Figs. S13, S14). As expected, a deconvolution model^45^ of the whole-blood expression of these seven plasma cell-related genes, adjusted for demographics, BMI, education level, ICS, OCS, and cell-type composition, confirmed their plasma cell-level association with the DNAm score (GALA II P=0.038, SAGE II P=7.5e-4; Supplementary Note S5).

*TNFRSF13B* and *TNFRSF17* encode key receptors that regulate B cell and plasma cell survival and differentiation^52,53^. Dysregulation of these pathways promotes mechanisms implicated in allergic asthma^62^ and autoimmunity^63^ associated with moderate-to-severe asthma, including persistent eosinophilia and exacerbations^64^. *SIGLEC10*, expressed across multiple immune cell populations, mediates immune suppression through interaction with soluble CD52; its altered expression has been linked to autoimmune responses and may contribute more broadly to immunomodulation in asthma^57,65–67^. *IGHV6-1* and *IGLV5-45* encode immunoglobulin variable chains characteristic of plasma cell antibody production, with *IGHV6-1* specifically implicated in allergic and autoimmune antibody repertoires^68,69^. *ITM2C*, *TXNDC5*, and *TXNDC11*, though less studied in asthma, are involved in the endoplasmic reticulum secretory machinery of plasma cells, coordinating protein folding, quality control, and trafficking to support high-rate antibody synthesis and secretion^70–72^. Notably, in both endothelial cells and multiple organ fibroblasts, *TXNDC5* also participates in hypoxia-responsive pathways involving TGF-β-driven fibrosis, redox adaptation, and NF-κB-mediated inflammation; all processes that may contribute to airway remodeling, chronic inflammation, and autoimmune features seen in a subset of severe asthma^73,74^. Collectively, these findings define a DNAm-associated sub-endotype marked by altered B-cell lineage gene expression within a T2-high context, which we term T2-high asthma with altered B cell response (T2ABC).

### T2ABC patients exhibit poor prognosis in a randomized clinical trial of omalizumab

Most approved biologic therapies for moderate to severe asthma target the T2 inflammatory pathway^10^. Therefore, to further evaluate the clinical relevance of the T2ABC sub-endotype, we asked whether T2ABC patients exhibit different clinical outcomes with T2-targeted biologic therapy. To investigate this, we retrospectively analyzed the EXTRA randomized controlled trial of omalizumab in T2-high asthma patients^15^. We focused on a subset of 300 participants with baseline (pre-treatment) whole-blood microarray gene expression data (151 treated with omalizumab and 149 placebo controls)^75^.

We first repeated the analysis reported in the original study. In the full trial sample (n=848), a Poisson regression analysis of 1-year asthma exacerbation rates—adjusting for baseline exacerbation count, baseline medication type, dose schedule, and time in the trial—reproduced the treatment effect reported by the authors (incidence rate ratio (IRR) of 0.76, 95% CI [0.61, 0.93]; P=0.0088). Analyzing the subset with available gene expression data (n=300) yielded a concordant effect estimate, though it was not statistically significant due to the reduced sample size (IRR=0.84, 95% CI [0.62, 1.15]; P=0.28; Supplementary Fig. S15).

To evaluate whether the T2ABC profile corresponds to differential prognosis, we computed a composite T2ABC score by summing the expression levels of the T2-high-specific DEGs available in the data (ten out of 12; Methods). Including this score as a covariate in the Poisson regression model revealed a 24% relative increase in the exacerbation rate per unit increase in the T2ABC score (IRR 1.24, 95% CI [1.07, 1.43]; P=0.004; Supplementary Fig. S16). A Cox proportional hazards analysis of time to first exacerbation—adjusting for the same covariates—yielded consistent results (hazard ratio (HR) of 1.22, 95% CI [1.03, 1.44]; P=0.021; Supplementary Fig. S17).

Next, we examined whether a subset of the ten genes in the composite T2ABC score contributed disproportionately to the prognostic signal. Repeating the Poisson regression analysis while replacing the composite score with the ten individual gene expression values identified *TXNDC5* as the only significant predictor (IRR 1.54, 95% CI [1.06, 2.24]; P=0.025; Supplementary Fig. S18). *TXNDC5* alone recapitulated the effect of the full T2ABC score on exacerbation rate (IRR 1.28, 95% CI [1.12, 1.47]; P=4.42e-4; Supplementary Fig. S19), including within the omalizumab-treated subgroup only (IRR 1.42, 95% CI [1.12, 1.78]; P=0.003; Supplementary Fig. S20). These results suggest that whole-blood *TXNDC5* expression can serve as a practical surrogate marker for T2ABC status.

Lastly, to assess whether T2ABC patients benefit from omalizumab despite their poor prognosis, we compared individuals in the upper quartile of *TXNDC5* expression (T2ABC-high) with those in the lower quartile (T2ABC-low). Responders were defined as patients whose 1-year exacerbation count decreased relative to baseline. As expected, T2ABC-low patients treated with omalizumab showed a significantly higher response rate than T2ABC-low patients in the placebo arm (Fisher’s exact test; P=0.019). In contrast, no treatment effect was observed among T2ABC-high patients (P=0.48), suggesting that this subgroup represents molecular variation orthogonal to omalizumab’s mechanism of action.

To explore the relevance of *TXNDC5* and the broader set of seven plasma cell-predominant T2ABC genes to omalizumab treatment resistance, we examined their expression in eight independent single-cell RNA-seq datasets (Methods). In both peripheral blood and intestinal tissue, these genes localized to highly secretory plasma cell subsets (Supplementary Fig. S21 and S22). Given the scarcity of IgE-producing plasma cells in circulation—approximately 0.3% of plasma cells in atopic patients with elevated IgE and approximately 1 of 100,000 circulating leukocytes^76^—detecting T2ABC patterns in whole-blood gene expression renders them likely associated with IgG- or IgA-secreting activity. To address the possibility that these patterns reflect aberrantly high frequencies of IgE-producing plasma cells, we examined total serum IgE levels in the EXTRA study and found no significant difference between T2ABC-high and T2ABC-low patients (Wilcoxon; P≥0.12, using either *TXNDC5* or the broader gene signature to define T2ABC-low/high). Omalizumab may therefore be less effective in T2ABC patients because their disease is not exclusively IgE-driven. Instead, it may involve immune complex formation, autoantibody-associated inflammation, or activation of eosinophils and neutrophils—all potentially mediated by IgG or IgA. While each of these pathways has been implicated in asthma pathogenesis through mechanisms unlikely to be affected by IgE blockade^64,77–83^, they are consistent with the T2ABC sub-endotype of T2-high asthma, as they may be nested within a broader T2 inflammatory context

## Discussion

Current asthma classification systems often fail to distinguish patients with similar biomarker profiles but divergent clinical outcomes^84^. The T2-high endotype, while foundational for molecular classification and therapeutic development, illustrates both the promise and limitations of biomarker-guided approaches^6^. Patients with ostensibly similar T2 profiles can experience markedly different disease trajectories, underscoring the need for more precise stratification^85^. DNAm offers a particularly compelling dimension for this purpose, as it integrates genetic predisposition, environmental exposures, developmental programming, and stochastic variation to modulate gene regulation^86–89^. Although DNAm changes may not be causal, they can mediate or be correlated with key molecular processes in asthma pathogenesis, thereby enabling finer resolution of disease heterogeneity. In contrast to static germline variants, DNAm signatures are dynamic—evolving across the life course and associated with disease states—yet exhibit greater stability than RNA-based measures^33,34^. This unique balance of responsiveness and reliability positions DNAm as an effective input for robust data-driven patient stratification and refinement of biomarker-guided therapeutic approaches.

In two racially and ethnically diverse pediatric asthma cohorts, we developed and applied PACA, a novel contrastive learning patient stratification method, to whole-blood DNAm data. The resulting DNAm score captured key indicators of clinical severity that routinely guide asthma management, including reduced FEV1/FVC ratios, elevated exacerbation burden, and increased oral corticosteroid use. Such hallmarks of high-risk asthma often go unrecognized until after prolonged symptom progression and higher healthcare utilization^10^. While our data reflect cross-sectional associations without establishing prospective predictive value, the DNAm score may serve as a real-time molecular marker of disease burden, offering a clinically actionable tool for stratifying patients prior to crisis-level decompensation, such as emergency department visits or hospitalization. These findings reinforce and expand upon growing evidence that omics-based profiling can provide clinically meaningful indicators of asthma phenotypes: DNAm and transcriptomic signatures from nasal, airway, and blood samples have been linked to lung function, BDR, symptom control, exacerbation risk, and overall severity^47,90,91^.

Among the clinical features routinely used to guide asthma management, biomarkers such as BEC and IgE are especially actionable, serving as key indicators of T2 inflammation and guiding biologic treatment selection^10,92^. Although both markers correlate with BDR in research settings, their predictive value is inconsistent and insufficient for clinical assessment of response to bronchodilators^92,93^. Our findings suggest this inconsistency reflects molecular heterogeneity that DNAm stratification can clarify. Across the DNAm score continuum, associations between BEC or IgE and BDR ranged from strong to absent, indicating that the clinical relevance of these biomarkers varies with underlying epigenetic context. This supports a shift in respiratory precision medicine, wherein DNAm scoring provides molecular context that enhances the individualized interpretation of traditional biomarkers such as BEC and IGE—clarifying T2 inflammation phenotypes and improving alignment with therapeutic decisions, including biologic eligibility.

To elucidate the biological basis of DNAm score-captured heterogeneity, we first examined cell-type-specific methylation patterns and identified a replicable eosinophil-specific hypermethylation signal at eight CpG sites, validated in an independent dataset of isolated eosinophils. This cell-type specificity indicates that, among individuals with similar T2-high profiles, epigenetic variation within eosinophils reflects distinct molecular programs–revealing functional diversity that may be associated with, or mechanistically involved in, disease expression and treatment response. Prior studies support this interpretation, showing that eosinophil methylation patterns vary by IgE level and clinical phenotype, further underscoring the biological heterogeneity embedded within the broader T2-high classification^46,49^. These epigenetic differences may reflect chronic cytokine exposure or divergent developmental and regulatory trajectories shaping eosinophil function^94,95^.

The DNAm score also stratified transcriptional activity within the T2-high endotype, with canonical markers such as *IL5RA*, *ST2*, and *CCR3* exhibiting a graded increase in expression consistent with escalating T2 inflammatory activity. In addition to these established pathways, we identified a replicable 12-gene transcriptional signature—designated T2-high asthma with altered B cell response (T2ABC)—associated with B-cell-lineage activity and likely missed in prior analyses using conventional statistical methods or transcriptomic data alone. T2ABC is orthogonal to conventional T2 definitions and represents a distinct molecular subtype within the T2-high endotype. This signature, which defines the T2ABC sub-endotype associated with greater disease severity and poor response to IgE-targeted therapy, likely reflects humoral immune activation marked by plasma cell differentiation. It encompasses B cell survival and differentiation signals (*TNFRSF13B* and *TNFRSF17* via BAFF/APRIL pathways, with *SIGLEC10* providing inhibitory modulation of activation), clonal expansion and immunoglobulin gene rearrangement (*IGHV6-1*, *IGLV5-45*), and upregulation of endoplasmic reticulum secretory machinery (*ITM2C*, *TXNDC5*, *TXNDC11*) to support elevated antibody production. This constellation of features is consistent with sustained plasma cell activity and enhanced antibody production.

To assess the clinical relevance of the T2ABC program, we tested its corresponding transcriptomic signature in an independent randomized trial of omalizumab. Despite active treatment, individuals with elevated expression of the signature—particularly *TXNDC5*—exhibited more frequent and earlier exacerbations, highlighting its potential to identify patients with treatment-resistant disease. Although the signature includes plasma cell-associated genes that are broadly expressed across isotypes, it is unlikely to solely reflect IgE-producing plasma cells. IgE levels alone do not explain treatment resistance, as omalizumab remains highly effective even among patients with markedly elevated IgE concentrations, well above those observed in the EXTRA dataset^15,96^. Furthermore, our analyses were conducted in peripheral blood, where plasma cells are rare; on average, IgE-producing plasma cells comprise only ∼0.3% of circulating plasma cells and ∼0.001% of total leukocytes^76^—too few to explain the observed gene expression patterns. Instead, the findings likely reflect increased antibody secretion of circulating IgA- and IgG-producing plasma cells, which together account for ∼90% of the peripheral plasma cell population^97^. This conclusion is supported by the observation that expression variation in the seven plasma cell-predominant genes of the T2ABC signature is driven by circulating plasma cells in both discovery and validation datasets, as well as in independent single-cell RNA-seq data.

Transcriptional signals of increased IgA- and IgG-producing plasma cell activity, together with eosinophil-specific hypermethylation patterns and unresponsiveness to IgE blockade, point to immunoglobulin-mediated mechanisms that contribute to asthma pathogenesis independently of classical T2 cellular pathways, yet within a persistent T2 inflammatory milieu. Both IgA and IgG can activate eosinophils, promoting degranulation and the recruitment of additional effector cells, thereby amplifying eosinophilic airway inflammation^98,99^. IgG immune complexes can further drive inflammation through complement activation and Fc gamma receptor (FcγR) signaling^99,100^. Emerging evidence also supports an autoimmune asthma subtype marked by IgG autoantibodies targeting endogenous immune proteins, including eosinophil granule components, which may amplify T2 inflammation and establish a self-sustaining, treatment-refractory loop^64,80–83^. This autoimmune program recapitulates key clinical and molecular features of T2ABC, including resistance to IgE blockade and the convergence of T2 inflammation with plasma cell-driven immune activity.

Unlike prior transcriptomic markers of biologic response, which have largely reflected T2 inflammation^101^, the 12-gene T2ABC signature is distinctly T2-orthogonal and derived from epigenetically defined patient stratification These features position the T2ABC signature as a clinically actionable biomarker that may help refine biologic prescribing and guide therapeutic development beyond the T2 paradigm. Moreover, because the PACA-derived DNAm score is trained and validated on diverse populations and independent of fixed biomarker thresholds, it may help address disparities in drug eligibility and improve equitable access to precision asthma therapies^102,103^. Replication across additional biologic agents, longitudinal assessment of methylation and expression dynamics, and mechanistic dissection to understand the underlying pathophysiology—as well as to identify actionable therapeutic targets—will be critical for widespread clinical translation.

While these findings offer a compelling framework for epigenetic stratification in asthma, several limitations warrant consideration to inform future refinement and application. First, the use of whole blood for methylation and gene expression profiling may not fully capture disease-relevant biology in the airway or lung tissues. Nevertheless, blood-based assays provide a scalable and clinically practical platform for population-level stratification, and the eosinophil- and B-cell-related signatures identified here align with known asthma immunopathology, supporting their biological plausibility^62,64,104^. Second, the cross-sectional design of our pediatric cohorts limits inference on the temporal dynamics of DNAm alterations in relation to disease progression and treatment response. Longitudinal studies that track methylation and transcriptional changes before, during, and after therapy will be essential to assess the durability and reversibility of these signatures. Third, biologic response analyses were retrospective and limited to a single agent, omalizumab; both prospective and retrospective evaluations across other targeted therapies—including anti-IL-5, anti-IL-5Rα, anti-IL-4Rα, and anti-TSLP agents—will be necessary to evaluate generalizability^10^.

The T2ABC signature distinguishes non-responders within T2-high asthma, however, its specificity outside canonical T2 inflammation remains uncertain. We cannot exclude the possibility that the presumed altered B-cell response captured by the signature is activated by alternative immune contexts and may influence treatment outcomes in ways not yet captured by available data. Addressing this question will require additional treatment-response datasets that include T2ABC-high individuals, alongside mechanistic studies to better delineate the boundaries of this sub-endotype. Additionally, given that the DNAm score reflects epigenetic variation alone, integrating into the stratification model complementary omic layers—such as transcriptomic, proteomic, and exposomic data—may further illuminate disease mechanisms and improve predictive accuracy. Finally, targeted functional studies will be necessary to define the roles of key T2ABC genes in airway inflammation, B cell regulation, plasma cell activity, and resistance to T2-directed biologics. These efforts will be critical to refine stratification-based discoveries and translate them into clinically actionable biomarkers and therapeutic strategies for severe asthma.

In summary, this study integrates methodological innovation, biological insight, and clinical relevance to advance molecular stratification in asthma. Methodologically, we developed PACA—a contrastive learning approach applied to whole-blood DNAm—to derive a robust, generalizable epigenetic score. Our DNAm score reveals meaningful heterogeneity within the T2-high endotype across four ancestrally and ethnically diverse cohorts spanning childhood and adulthood. Biologically, the score captures eosinophil-specific hypermethylation and unmasks a distinct immune program marked by altered B cell response (T2ABC). Clinically, it is associated with markers of asthma morbidity and yields a transcriptomic signature prognostic of poor outcomes despite biologic treatment. Collectively, these findings demonstrate that DNAm-based stratification can refine interpretation of conventional biomarkers and expose previously unrecognized disease mechanisms. More broadly, PACA offers a scalable framework for identifying clinically actionable hidden subtypes across heterogeneous diseases.

## Data Availability

Data produced in the present study will be made available after formal peer-review publication

## Methods

### Primary study populations

#### GALA II and SAGE II

Clinical, biomarker, spirometric, DNAm, and gene expression data were collected as part of GALA II and SAGE II, two case-control studies of asthma conducted between 2006 and 2014. GALA II enrolled participants from five urban study centers across the United States (Chicago, Bronx, Houston, San Francisco Bay Area, and Puerto Rico), while SAGE II recruited participants exclusively from the San Francisco Bay Area. Participants, self-identifying as African American with four African American grandparents in SAGE II and as Latinos in GALA II, ranged from 8 to 21 years at recruitment. Case participants were defined by physician-diagnosed asthma, presence of asthma symptoms, or use of asthma medication within the last 2 years, excluding any history of other lung or chronic nonallergic illnesses. Healthy control participants exhibited no lifetime history of asthma or allergies and reported no instances of coughing, wheezing, or shortness of breath in the two years preceding enrollment. Case and control participants were matched at a 1:1 ratio based on age within one year at each recruitment site. The inclusion and exclusion criteria have been described in detail elsewhere [1, 2]. The University of California San Francisco Human Research Protection Program Institutional Review Board approved study protocols for GALA II and SAGE II.

#### Epigenome-wide methylation assessment and quality control

Samples for DNAm assessment were collected at the same time when spirometric measurements and BDR were measured. Genomic DNA was extracted from whole blood using Wizard Genomic DNA Purification Kits (Promega, Fitchburg, WI). DNAm measurements were obtained at more than 850,000 CpG sites from whole blood using the Illumina Infinium MethylationEPIC array (Illumina, San Diego, CA). Quality control was conducted using the ENmix R package (version 1.22.0), involving background and dye bias correction, probe-type bias adjustment, inter-array normalization, and the removal of low-quality CpG probes and samples [3]. We excluded probes from sex chromosomes and probes that are either polymorphic or cross-reactive [4, 5]. Methylation intensities were used to calculate beta values, and outlier or missing DNAm data points were replaced by imputed values unless the probe or sample missingness rate exceeded 5% and 10%, respectively. See additional preprocessing information in Supplementary Note S3.1.

#### RNA sequencing data

Whole-transcriptome data generated from whole blood were sequenced as part of the Trans-Omics for Precision Medicine (TOPMed) WGS program [6]. Processed whole-blood RNA-seq data were available for a subset of asthma patients with DNAm profiles in the GALA II and SAGE II cohorts. Among participants with paired data (n=774; across GALA II and SAGE II), 92.5% (n=716) had RNA-seq and DNAm derived from the same biospecimen. In 7.0% (n=54), the two data types were obtained from the same individual at different time points, while sample provenance was unknown for the remaining 0.5% (n=4).

Processed RNA-seq data were obtained from the study by Kachuri et al. [7], which provides full details on experimental protocols, data processing, and quality control procedures. Briefly, total RNA was extracted from PAXgene blood samples, depleted of globin, and assessed for quality before undergoing automated library preparation using a strand-specific Illumina TruSeq protocol, followed by sequencing on the HiSeq 4000 platform targeting 50 million paired-end reads per sample. Reads were processed using the TOPMed RNA-seq pipeline with GRCh38 and GENCODE 30, and count-level data were quality-controlled and normalized per GTEx v8 protocols [8].

#### Biomarker, spirometric, and BDR assessments

Serum total IgE levels were measured twice using the Phadia 100 system (ThermoFisher Scientific in Uppsala, Sweden). If the results were not within 10% agreement, a third measurement was taken. We applied log transformation to the serum total IgE values to address the right-skewed distribution of the data on its natural scale. Peripheral BECs were obtained from complete blood cell counts (CBC) with differentials from certified laboratories. Quest Diagnostics was used for GALA, while the University of California San Francisco Clinical Laboratories was used for SAGE II.

Spirometric measurements were obtained from all participants both before and 15 minutes after the administration of four puffs of albuterol (90 µg per puff), following the American Thoracic Society recommendations [9]. Spirometry was conducted for a third time following a second albuterol dosage (two puffs for those <16 years old or four puffs for those ≥16 years old). Spirometry and bronchodilator testing were conducted using a KoKo® PFT Spirometer (nSpire Health Inc., Louisville, CO). All asthma medications were held for 12 hours prior to the spirometry assessment. Pulmonary function percent predicted (observed⇥100/predicted) baseline and z-scores were calculated using the Global Lung Function Initiative race-neutral equations [10]. BDR was determined by assessing the percentage change in measured FEV1 before and after albuterol administration, utilizing post-albuterol spirometry with the greatest observed change. Positive BDR (response) was defined as an increase in FEV1 from baseline equal to or exceeding 12% predicted. BDR was evaluated concurrently with the collection of whole-blood samples used for collecting blood biomarkers and DNAm profiling.

#### Imputing missing blood eosinophil counts

BEC and CBC measurements are only available for 255 (37.2%) of the GALA II and 83 (19.3%) of the SAGE II cases. To impute eosinophil proportions for samples with missing BEC levels, we applied the cell-type decomposition approach described in the method BayesCCE [11] (“impute mode”) as follows. First, we leveraged the available CBC measurements from a subset of the GALA II cases and the corresponding whole-blood DNAm profiles of these samples to estimate cell-type specific DNAm signature profiles. We considered cell-type signatures of 450 CpGs previously identified to be informative for leukocyte mixture decomposition [12]. Samples with abnormal measurements, defined as those with more than 2% deviation between their whole-blood counts (WBC) and the sum of their neutrophil, eosinophil, lymphocyte, monocyte, and basophil absolute counts, or those with reported eosinophil cell-type proportions exceeding 30%, were excluded from the step of estimating signature profiles. Given cell-type DNAm profiles estimated from the known cell compositions, we imputed missing cell-type proportions for participants from both cohorts using quadratic programming [11].

#### Covariates and asthma phenotypes

In all linear regression models, including the interaction models, we accounted for age, sex, BMI, fractions of African and European genetic ancestry (Supplementary Note S3), Mexican ethnicity (indicator; self-reported), education level (on an ordinal scale; self-reported), medication use frequency in the last 2 weeks (self-reported), ICS use (self-reported), and OCS use in the last month and year (self-reported). In any analysis combining both GALA II and SAGE II, we also accounted for the cohort (a binary indicator variable). In any given analysis, we considered only samples with no missing covariates or phenotypes. Further information and description of asthma phenotypes, including exacerbation count, hospital visits, and emergency room visits in the last year, are provided in Supplementary Note S3.

### PACA: Phenotype Aware Component Analysis

#### Problem setup

We consider a low-rank model for high-dimensional data coming from a target population and a background population. Both populations share the same biological and non-biological sources of variation, with the exception of an additional component in the target population that represents phenotypic heterogeneity. Hereafter, we refer to the target and background groups as “cases” and “controls”, respectively.

Let 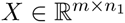 be a matrix of measurements of m features for *n*_1_ cases, and let 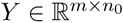 be a matrix of measurements of the same m features for *n*_0_ control individuals, such that *m* > *max*(*n*_0_, *n*_1_). We consider the following descriptive model:

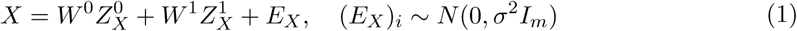

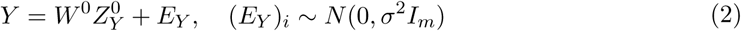

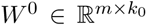 denotes the sources of variation (directions in the features space ℝ*^m^*) of a *k*_0_-dimensional low-rank signal (typically *k*_0_ << *min*(*n*_0_, *n*_1_)), and 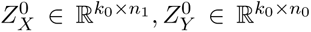 denote components (in the sample spaces 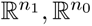) of individual-level variation for the individuals in *X*, *Y*, respectively. Altogether, these directions and components represent the shared variation across cases and controls. 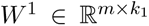 and 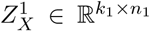 denote the sources of variation of an additional *k*_1_-dimensional low-rank signal and its corresponding individual-level component, representing the phenotypic heterogeneity of the cases in 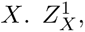 by construction, represents variation specific to cases and is where we expect to find variation in disease. Therefore, our goal is to learn 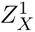 (up to a linear transformation). Of note, 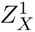 may be more subtle and explain much less of the variation in the data compared to 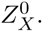

#### Orthogonality assumption

We make the assumption that *W* ^0^ ⊥ *W* ^1^. That is, we assume the case-specific sources of variation are orthogonal to the axes of common variation across cases and controls. We refer to this assumption as the orthogonality assumption. Neglecting this assumption, while not incorporating further supervision, may lead to under-correction for shared sources of variation across cases and controls. This, in turn, can lead to falsely tagging background variation as phenotypic heterogeneity (Supplementary Fig. S5).

Under Eq. (1)-(2) and the orthogonality assumption, learning case-specific variation amounts to finding sources of variation in *X* that do not exist in *Y*. However, one should not necessarily remove from *X* all the variation shared across cases and controls for identifying case-specific variation, as this may be suboptimal. For instance, in cPCA [13], a contrastive method similar in spirit to PACA, incorporating the orthogonality assumption by setting a very large contrastive hyperparameter is suboptimal (Supplementary Note S1.4). An arguably better approach is to remove only sources of variation that exist in both *X*, *Y* and are more dominant in the data compared to the case-specific variation. Such sources of variation are expected to be of much lower dimension than the dimension of the observed data. This is exactly the key idea behind PACA.

#### The PACA algorithm

Using a singular value decomposition we can consider an alternative formulation for the model in (1)-(2):

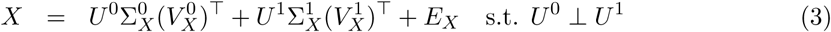

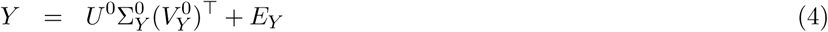

where each of 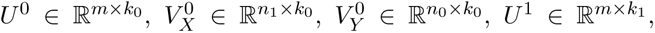 and 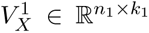 represents an orthonormal basis. Under this presentation, we are interested in learning 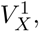, a (possibly linearly-transformed) surrogate for 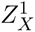 in Eq. (1). Given the directions of the shared sources of variation *U* ^0^, note that

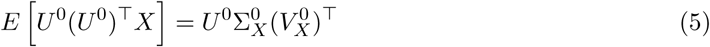

which establishes a way to remove the expected effect it induces on *X* (i.e., the signals in *X* that are coming from sources of variation that exist in both cases and controls). Our PACA algorithm, therefore, estimates and removes the effects of *U* ^0^ on *X*, followed by PCA for capturing the low-rank structure 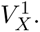 Our main focus is thus estimating *U* ^0^.

Since *U* ^0^ is found in both *X*, *Y*, we can estimate it by employing a canonical correlation analysis (CCA) [14]. Importantly, unlike in a typical application of CCA, where we wish to find linear transformations of the features that yield the highest correlation between the same set of samples in two datasets (i.e., vectors in the samples space), here, we seek linear transformations of the samples that provide vectors in the features space ℝ*^m^*. Specifically, we find the first pair of canonical variables by solving:

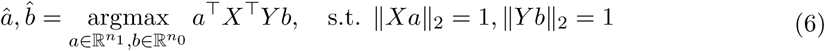

Setting 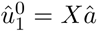 yields the representation in *X* of the strongest direction of shared variation across *X* and *Y*. Let *S_XY_*, *S_XX_* and *S_YY_*, be the empirical sample cross-covariance and sample covariance matrices of *X* and *Y*. The solution for â, b^ is known to be given by the eigenvector that corresponds to the top eigenvalue of 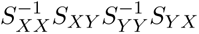 and the eigenvector that corresponds to the top eigenvalue of 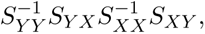 respectively [14, 15]. This procedure can be repeated iteratively by restricting 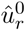 to be orthogonal to the previous canonical variables 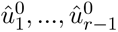 (and similarly for the variables of *Y*). Eventually, the collective of these vectors 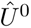 can be used for removing the shared sources of variation from X following Eq. (5); see a summary of PACA in Algorithm 1.

The number of canonical directions representing sources of variation shared between cases and controls is unknown. Accounting for too few canonical directions would result in PACA capturing variation unrelated to the heterogeneity of the condition of interest. Accounting for too many canonical directions, on the other hand, is expected to lead to power loss. We, therefore, employ a permutation scheme to identify the minimal number of canonical directions that need to be removed to identify significant case-specific variation (Supplementary Algorithm S.A.1; Supplementary Note S1.1).

##### Algorithm 1 Phenotype Aware Components Analysis (PACA)

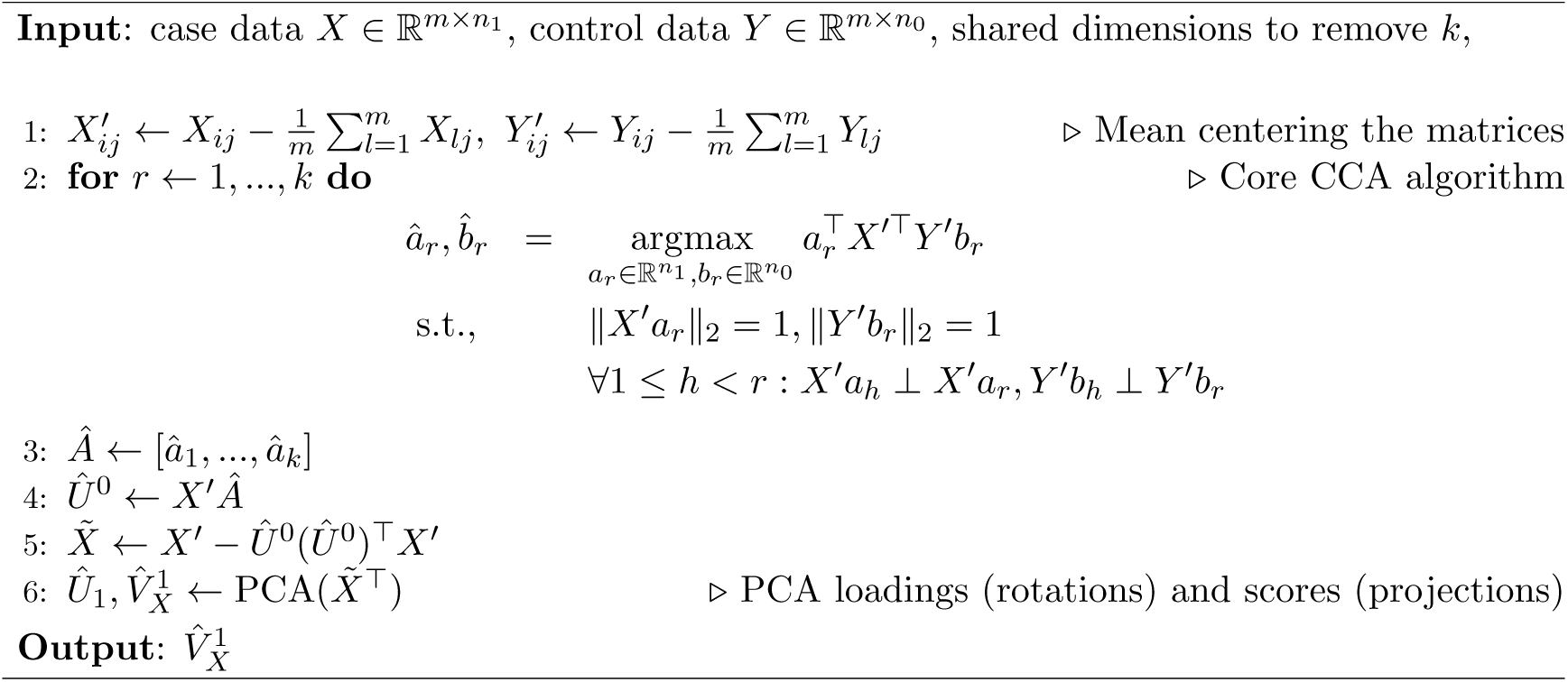

Lastly, PACA is constrained by the same limitations as a standard CCA, which requires, in our setup, more features than samples. While this is not a limitation in our study, as the number of CpGs used for developing the DNAm score is larger than the number of samples, to address scenarios where this requirement is unmet, we developed a randomized extension of PACA (rPACA). This randomized algorithm operates in the regime of more samples than features by estimating the PACA components via random data sampling (Supplementary Algorithm S.A.2; Supplementary Note S1.2).

#### Benchmarking PACA with alternative contrastive methods

We performed comprhensive benchmarking using synthetic data, as well as real DNA methylation, gene expression, and genotype datasets, to compare PACA with standard PCA, contrastiveVAE (cVAE) [16], and contrastive PCA (cPCA) [13]. While other contrastive learning methods exist (e.g., Sim-CLR [17]), they apply contrastive principles during model training to learn representation models, rather than *contrastive* representation models – patterns that are more variable in cases than in controls. We excluded such methods from our benchmarking because their objectives are not aligned with the specific goal of case-specific variation discovery. See Supplementary Note S2 for additional details and benchmarking results.

### Primary statistical analysis

#### Constructing DNAm score for asthma patient stratification

To identify asthma heterogeneity in DNAm data, we first created a panel of CpGs enriched for association with asthma. We anticipated that proper feature selection would improve performance by reducing noise and narrowing the search space. We used TCA [18] to perform an epigenome-wide association study (EWAS) with asthma status in GALA II (R package TCA, version 1.2.1). We accounted for age, sex, the top five genetic principal components (PCs), and the top 30 PCs calculated from the 50,000 lowest variance probes (treated as control probes; as previously suggested to account for technical variation [18, 19]). Since TCA requires cell-type proportions as an input, we applied EpiDISH [20] to estimate proportions using the cent12CT.m reference panel [21]. This analysis resulted in a panel of 7,662 CpGs passing a nominal association threshold of p<0.01, which we retained for constructing the PACA DNAm score.

We learned the PACA components using the set of 7,662 CpGs in GALA II (discovery) and applied the model to SAGE II (replication) by multiplying the coefficients of the PACA components by the same CpGs in SAGE. To evaluate whether the PACA components capture replicable asthma heterogeneity, we tested their linear correlation with a set of 668 phenotypes. Evaluating the consistency of these correlations across GALA II and SAGE II revealed poor consistency for the first PACA component and high consistency for the second PACA component (Supplementary Tables S3 and S4). We, therefore, set the second PACA component as the DNAm stratification score.

#### Testing BEC and IgE as differential biomarkers of drug response

Testing the statistical interaction effect of the DNAm score with BEC and IgE on BDR was based on linear regression models with BDR as the outcome. An interaction was evaluated by including in the regression a multiplicative term between the DNAm score and a given biomarker, in addition to a linear term of the biomarker and the addition covariates described earlier.

Restricting the statistical interaction analysis to T2-high patients was based on elevated BEC fractions (≥4%) or elevated IgE (≥100 kU/L). We also considered five other definitions of a T2-high status. Specifically, based on serum total IgE values only, dichotomized into high and low categories using 100 kU/L as the cut-off [22]; elevated eosinophil counts as defined by peripheral BEC values based on 150 c/µL and 300 c/µL cut-offs [23]; and imputed BEC proportion values, binarized into high or low based on a 4% cut-off [24, 25]. Testing the statistical interactions among patients with low baseline lung function was restricted to patients with a cut-off of 80% predicted percent FEV1 or a cut-off of 0.70 for the FEV1/FVC ratio [26]. Finally, reported odds ratios were calculated based on unadjusted logistic regression models with the response status (BDR≥12%) as the outcome.

#### Benchmarking predictive models of BDR

Three distinct model types were assessed, including a model based on DNAm levels, biomarker-based models, and a model based on cell-type composition. In all models, we used the GALA II cohort as the training set and the SAGE II cohort as the test set. The DNAm model utilized the same 7,662 CpGs used in our PACA-derived DNAm score as features. These features were used in an elastic net penalized logistic regression model, trained using a 10-fold cross-validation procedure to determine the optimal lambda parameter (R’s glmnet; default *α* = 0.5 ratio for the *l*_1_ and *l*_2_ penalties). For the biomarker-based models, we considered a logistic regression model, incorporating log-transformed total IgE, imputed BEC, and their interaction as predictors; some of the models considered only subsets of patients based on different percentile groups of the DNAm scores. The cell-type composition model was constructed using logistic regression with imputed fractions of lymphocytes, monocytes, basophils, neutrophils, and eosinophils as the predictors. Model performance was evaluated using the area under the receiver operating characteristic curve (AUC-ROC) and the area under the precision-recall curve (AUC-PR).

#### Cell-type-level differential methylation

We used TCA [18] to identify cell-type-level differential methylation with the DNAm score. As input to TCA, we provided the bulk DNAm levels of the top 100 most informative features of the DNAm score model – ranked by the absolute value of their coefficients – that mapped to at least one gene based on the Illumina MethylationEPIC manifest file (v1.0 B4). TCA models differential methylation in cell types for which per-sample proportions are available. Therefore, similarly to our application of TCA for feature selection before applying PACA, we used Epidish [20] and a methylation reference panel [21] to estimate cell-type proportions for a set of 10 immune cell types, including neutrophils, eosinophils, monocytes, CD4 näıve T, CD4 memory T, CD8 näıve T, CD8 memory T, B memory, B näıve, and natural killer cells.

To test for cell-type-level differential methylation in T2-high and T2-low patients, we used the same BEC and IgE cutoffs described earlier. Since our data include more T2-high (477 in GALA and 341 in SAGE) than T2-low samples (124 in GALA and 87 in SAGE), for the T2-low-specific differential methylation analysis, we increased statistical power by pooling samples across GALA II and SAGE II. TCA automatically accounts for cell-type composition using the cell-type proportions provided as input. In addition, in all models, we accounted for age, sex, the top five genetic PCs, and the top 10 PCs calculated from low-variance probes, calculated as described earlier. Of note, we considered only 10 PCs from low-variance probes (and not 30, as described earlier) due to the smaller sample size in this analysis.

#### Differential gene expression analysis

Genes were first filtered to retain those with variance between 0.001 and 2 on variance-stabilizing transformed (VST) normalized counts, calculated separately for each cohort and taking the intersection of qualifying genes across cohorts (34,076 genes; iso-forms included). The differential gene expression analysis was performed using DESeq2 (version 1.42.0) [27], while accounting for unknown confounders using surrogate variable analysis (SVA) [28] (sva R package).

For a given analysis, we defined a full model including the variable of interest and a reduced null model excluding it. In each model, we accounted for 10 SVA surrogate variables, age, sex, BMI, education level, African and European genetic ancestry proportions, ICS use, and OCS use. For GALA II, we included Mexican ethnicity as an additional covariate. We performed the analysis in GALA II and SAGE II separately.

#### Evaluating T2ABC genes using single-cell RNAseq data

To evaluate the cell-type-level expression of the 12 T2ABC genes, we used two annotated and curated CellHint organ reference maps [29] from the CellTypist [30] web portal. The human blood CellHint model (version v1), which integrates data from four adult human blood datasets [30–33], and the human intestine CellHint model (version v1), which integrates data from four adult human intestine datasets [34–37]. We further confirmed the expression of the seven genes predominantly expressed in plasma cells using data from the ImmGen project (data not shown) [38].

We used plasma cells from the CellHint blood (493 cells) and intestine (46,348 cells) models to construct plasma cell-specific single-cell reference maps. For each tissue, we applied Scanpy [39] to compute a UMAP embedding based on PCA of the top 2,000 most variable genes. To annotate secretory states within these maps, we quantified the combined expression of PRDM1, IRF4, and XBP1–key transcriptional regulators of plasma cell differentiation and antibody secretion, and central components of the unfolded protein response and secretory machinery.

#### Computational deconvolution of the seven plasma cell-related genes

We deconvolved the whole-blood gene expression levels of TNFRSF13B, TNFRSF17, IGHV6-1, IGLV5-45, ITM2C, TXNDC5, and TXNDC11 in GALA II, SAGE II, and the EXTRA datasets. For each dataset, we applied CIBERSORTx [40], a reference-based bulk RNA deconvolution method, together with the immune cell reference panel LM22 [41], which includes reference data for deconvolving plasma cell expression levels and proportions. To identify the cellular sources of each of the seven genes, we used the cell-type-level p-values returned by CIBERSORTx. To confirm the association between the plasma-cell-level expression of these seven genes and the DNAm score, we utilized the deconvolution model TCA [18], which enabled us to use the genes’ whole-blood expression to identify cell-type-level statistical effects on the DNAm score. Since TCA requires cell-type proportions as an input, we used the estimates provided by CIBER-SORTx [40]. See Supplementary Note S5 for complete details.

### Validating the eosinophil-specific differential methylation

#### Study population and isolation of human eosinophils

We used isolated eosinophil DNAm data from a subset of the SLSJ asthma family cohort of 1,394 individuals distributed in 271 independent families from a French-Canadian founder population [42]. Human eosinophils were isolated as previously described [43] from blood samples of a subset of 24 individuals from the SLSJ cohort, including 16 asthmatic cases and eight non-asthmatic controls. Briefly, after removing platelet-rich plasma with centrifugation of the 200 ml blood samples, erythrocytes and mononuclear cells were removed using Dextran-mediated sedimentation and lymphocyte separation medium, respectively. Remaining erythrocytes were removed using hypotonic lysis and other granulocytes were removed using negative selection with anti-CD16 MicroBeads. DNA was extracted using the QIAamp DNA Blood Mini Kit.

#### Methylation data

DNAm was assessed using Illumina Infinium HumanMethylation450 BeadChip arrays. Methylation levels were read and filtered using the RnBeads v2.18.1 R package [44, 45]. Normalization was done using the swan method [46], and background variation was removed using the methylumi.noob method [47], both implemented in RnBeads.

#### Validation analysis

We computed a composite hypermethylation score for each sample by summing the eosinophil DNAm levels of the three CpGs that overlapped between the Human-Methylation450 array and the nine CpGs identified in the MethylationEPIC-based differential methylation analysis using GALA II and SAGE II. Following the association between the DNAm score and pulmonary function in GALA II and SAGE II, we tested the eosinophil-specific composite score for association with pulmonary function in the SLSJ cohort (measured before and after albuterol administration), including FEV1 and FVC (percent predicted) and their ratio. Given the small sample size (n=24), both cases and controls were used, and statistical significance was assessed using a permutation test, with the linear correlation between the (permuted or unpermuted) variable and the composite score used as the test statistic.

### Validation using randomized controlled trial with omalizumab

#### Study population

We used data from the EXTRA study, a 48-week multicenter phase III, randomized, double-blind, placebo-controlled trial of the anti-IgE omalizumab efficacy for severe allergic asthma [48]. We leveraged whole-blood RNA collected in PAXgene tubes (microarray) at week 0 (pre-treatment baseline) and clinical phenotypes from participants in this study. Among 848 participants with uncontrolled asthma despite high-dose of ICS and long-acting *β*-agonists, 427 were randomized to omalizumab and 421 to placebo. Whole-blood microarray gene expression was later collected for a biomarker subcohort of 300 participants (151 omalizumab, 149 placebo) who had both PAXgene RNA and array data passing quality control metrics [49].

#### Clinical outcomes

The primary clinical outcome, asthma exacerbation rate, was defined as the annualized number of study-defined exacerbation requiring systemic corticosteroid treatment, emergency department visits, or hospitalizations. Time to first protocol-defined exacerbation was used as a secondary outcome for survival analysis. Treatment response was defined as a binary response variable, indicating whether there was a reduction in exacerbation rate compared to baseline.

#### T2ABC gene expression score

We constructed a T2ABC score by summing the standardized (z-score normalized) expression values of the available genes from the T2ABC 12-gene signature identified in GALA II and SAGE II (ten out of the 12 genes; IGHV6-1 and CEP85 were not available in the EXTRA data). Eventually, we standardized the T2ABC score across all samples to facilitate interpretation of effect sizes.

#### Poisson and Cox regression models for clinical outcomes

We employed Poisson regression with over-dispersion adjustment (quasi-Poisson family) to model the relationship between the T2ABC score and annualized exacerbation rates. In all Poisson models, study time was included as a normalized offset term – log-transformed study days divided by 365 – to compute annualized incidence rate ratios (IRRs). Cox proportional hazards regression [50] was used to model time-to-first-exacerbation, with hazard ratios (HRs) and 95% confidence intervals; this analysis was done using coxph function in the survival R package (version 3.5.5). Unless indicated otherwise, all regression models included the following covariates: dose schedule (2-week vs 4-week intervals), baseline exacerbation count in the previous year, baseline medication category (ICS/LABA, ICS/LABA with additional controllers excluding oral corticosteroids, or ICS/LABA with oral corticosteroids or ≥4 exacerbations), and treatment assignment (omalizumab vs placebo).

## Code availability

All statistical analyses were performed with R v.4.3.2 (R Foundation for Statistical Computing, Vienna, Austria). We provide a highly efficient implementation of PACA in C++ using the Eigen library with an wrapper R package for a user-friendly interface. The PACA package is available via GitHub at github.com/adigorla/PACA.

## Acknowledgements

A.G., E.R. and J.F. were supported, in part, by the National Institute of Mental Health grant R01MH122569. E.R. was supported by the National Human Genome Research Institute grant 1R21HG013393. A.G and J.F were also supported, in part, by the National Institute of Mental Health grant R01MH130581. J.P.G. was supported by fellowship FPU19/02175 (Formacíon de Profesorado Universitario Program) from the Spanish Ministry of Science, Innovation, and Universities (MICIU). This work was supported in part by the Sandler Family Foundation, the American Asthma Foundation, the RWJF Amos Medical Faculty Development Program, Harry Wm. and Diana V. Hind Distinguished Professor in Pharmaceutical Sciences II, the American Lung Association [CAALA2023], the National Institutes of Health, National Heart, Lung, and Blood Institute [Grants 1K23HL169911, R01HL155024-01, R01HL117004, R01HL128439, R01HL135156, X01HL134589], the National Institute of Health and Environmental Health Sciences [R01ES015794, R21ES24844], the National Institute on Minority Health and Health Disparities [R01MD010443, R56MD013312], and the Tobacco-Related Disease Research Program [24RT-0025, 27IR-0030]. This research was also supported by the UK Biobank Resource under application 33127.

The authors thank the participants and their families for their contribution, as well as the health care professionals and clinics for their support and participation in the Genes-Environments and Admixture in Latino Americans Study and the Study of African Americans, Asthma, Genes and Environments. In particular, the authors thank the study coordinator Sandra Salazar, the African American Wellness Project (AAWP) for supporting our community engagement and the recruiters who obtained the data: Kelley Meade, MD, Adam Davis, MPH, MA, Lisa Caine, Emerita Brigino-Buenaventura, MD, Duanny Alva, MD, Gaby Ayala-Rodriguez, Ulysses Burley, Elizabeth Castellanos, Jaime Colon, Denise DeJesus, Iliana Flexas, Blanca Lopez, Brenda Lopez, MD, Louis Martos, Vivian Medina, Juana Olivo, Mario Peralta, Esther Pomares, MD, Jihan Quraishi, Johanna Rodriguez, Shahdad Saeedi, Dean Soto, Ana Taveras, and Emmanuel Viera.

The generation of molecular data for the TOPMed program was supported by the National Heart, Lung, and Blood Institute (NHLBI). RNA-seq for the NHLBI TOPMed Genes-Environments and Admixture in Latino Asthmatics Study (GALA II; phs000920) and Study of African Americans, Asthma, Genes, and Environments (SAGE; phs000921) was performed at the Broad Institute Genomics Platform (HHSN268201600034I). We also thank the participants of UK Biobank and the support from the Hoffman2 HPC administrators at UCLA.

