## Supplementary Materials for "Epigenetic patient stratification reveals a sub-endotype of type 2 asthma with altered B-cell response"

### Part I

### Supplementary Notes

**S4 Alternative models for predicting bronchodilator response** **17**

**S5 Computational deconvolution of the seven plasma cell-related genes extended details** **18**

### S1 Phenotype Aware Component Analysis extended details

#### S1.1 Estimating the dimension of the shared variation

In typical settings, the parameter $k_0$, which represents the dimension of $U^0$, is unknown. We may estimate it as the largest value $r$ that reveals a significant correlation between the correlated subspace of $X$ and $Y$ induced by $\hat{a}_r, \hat{b}_r$; this can be achieved, for example using Bartlett's Chi-squared test or Rao's approximate F statistic [1]. However, in practice, this approach could lead to over-regularization of the data by removing too many axes of shared variation. In order to see why, recall that our goal is to be able to systematically target variation of interest (i.e., disease heterogeneity) that may be weaker than the dominant sources of variation in the data. Standard clustering and dimensionality reduction tools are unlikely to achieve this when applied to the original data. Yet, we expect them to be effective in revealing the target variation in $X$ if we apply them to $\tilde{X}$, a residualized (adjusted) version of the data, in which all sources of variation stronger than the target variation are removed. In other words, there is no need to estimate the true $k_0$ and remove a structure of this dimension from the data prior to applying dimensionality reduction. Instead, we wish to find what is the minimal number of axes of variation $k$ that we need to remove in order to reveal variation that is unique to cases. Below, we provide a brief description of the algorithm for selecting the dimension $k$; see Algorithm S.A.1 for complete details.

Given all axes of shared variation learned from the data using the procedure in Eq. (6), we learn $k \in \{1, ..., \min(n_0, n_1)\}$ using binary search as follows. Given a candidate parameter $k$, we evaluate the variance of the top PC $\hat{z}$ we calculate from $\tilde{X}$, the residualized $X$ accounting for the top $k$ axes of shared variation; this same PC, which may reflect case-specific variation, is also used for evaluating the variance it captures in $\tilde{Y}$ – the residualized $Y$ matrix – upon projection. Comparing these variances to "null" variances obtained by permuting the loadings of $\hat{z}$ allows us to determine whether accounting for $k$ axes of shared direction is sufficient to detect signal that is unique to $X$.

This evaluation can lead to one of four scenarios, based on which we decide on the next partition of the binary search: there can be (i) significant variation in cases but not in controls, (ii) significant variation in cases and controls, (iii) no variation in cases or controls, or (iv) significant variation in controls but not in cases. Scenarios (i) and (iii) lead us to consider lower values of $k$ due to possible over-correction. The former means we revealed case-specific variation, yet, we may be able to refine the signal if we can identify it using lower $k$, and the latter indicates that all the structure in the data has been removed. Scenario (iv) indicates a violation of our model assumption, which leads to termination, and lastly, scenario (ii) indicates residual shared variation, which suggests increasing $k$.

Empirically, we found that considering a threshold $\Delta$ on the maximum ratio between the variance $\hat{z}$ explains in $\tilde{X}$ and the variance it explains in$\tilde{Y}$ is effective in discerning between the different scenarios. For example, if the ratio between the variances of $\tilde{X}$ and $\tilde{Y}$ is greater than $\Delta$ then we admit scenario (i). In practice, we found that $\Delta = 10$ is a reasonable choice and is the default in all analyses in this study.

**Algorithm S.A.1** PACA: selecting $k$ (the dimension of shared variation to remove)

**Input**: $X \in \mathbb{R}^{m \times n_1}$, $Y \in \mathbb{R}^{m \times n_0}$, $\Delta$ variance ratio threshold, $\gamma \in [0,1]$ significance level

$X'_{ij} \leftarrow X_{ij} - \frac{1}{m}\sum_{l=1}^{m} X_{lj},\ Y'_{ij} \leftarrow Y_{ij} - \frac{1}{m}\sum_{l=1}^{m} Y_{lj}$ ▷ Mean centering the matrices

$k_{\min} \leftarrow 1, \quad k_{\max} \leftarrow \min(n_0, n_1), \quad d \leftarrow k_{\max}, \quad n \leftarrow 2\log_2(d)/\gamma$

**for** $r \leftarrow 1, ..., d$ **do**

$$\begin{aligned} \hat{a}_r, \hat{b}_r \quad &= \underset{a_r \in \mathbb{R}^{n_1}, b_r \in \mathbb{R}^{n_0}}{\operatorname{argmax}} \quad a_r^\top X'^\top Y' b_r \\ \text{s.t.,} \quad & \|X' a_r\|_2 = 1, \|Y' b_r\|_2 = 1 \\ & \forall 1 \leq h < r : X' a_h \perp X' a_r, Y' b_h \perp Y' b_r \end{aligned}$$

$A \leftarrow [\hat{a}_1, ..., \hat{a}_d], \quad B \leftarrow \left[\hat{b}_1, ..., \hat{b}_d\right]$

**while** $k_{\min} < k_{\max}$ **do** ▷ Binary Search

  $k \leftarrow (k_{\min} + k_{\max})/2$

  $\hat{U}_X^0 \leftarrow X_1 A_{1:k},\ \hat{U}_Y^0 \leftarrow Y_1 B_{1:k}$ ▷ $A_{1:k}, B_{1:k}$ indicate the first $k$ columns of $A, B$

  $\tilde{X} \leftarrow X' - \hat{U}_X^0 (\hat{U}_X^0)^\top X',\ \tilde{Y} \leftarrow Y' - \hat{U}_Y^0 (\hat{U}_Y^0)^\top Y'$

  $\hat{z}, \hat{v} \leftarrow \text{PCA}(\tilde{X}^\top)$ ▷ Top PC loadings (rotation) and scores (projection)

  $\text{counter}_X \leftarrow 0, \quad \text{counter}_Y \leftarrow 0$

  **for** $i \leftarrow 1, \ldots, n$ **do**

    $\hat{z}_{\text{perm}} \leftarrow \text{permute}(\hat{z})$ ▷ Permute the loadings of $\hat{z}$

    **if** $\text{Var}[\tilde{X}^\top \hat{z}_{\text{perm}}] > \text{Var}[\hat{v}]$ **then**

      $\text{counter}_X \leftarrow \text{counter}_X + 1$

    **if** $\text{Var}[\tilde{Y}^\top \hat{z}_{\text{perm}}] > \text{Var}[\tilde{Y}^\top \hat{z}]$ **then**

      $\text{counter}_Y \leftarrow \text{counter}_Y + 1$

  **if** $\text{counter}_X < 1 \land \text{counter}_Y \geq 1$ **then** ▷ Scenario (i) : Variation in cases but NOT in controls

    $k_{\max} \leftarrow k$

  **else if** $\text{counter}_X < 1 \land \text{counter}_Y < 1$ **then** ▷ Scenario (ii) : Variation in cases AND controls

    **if** $\text{Var}[\hat{v}] > \Delta \cdot \text{Var}[\tilde{Y}^\top \hat{z}]$ **then** ▷ Variation in cases > controls

      $k_{\max} \leftarrow k$

    **else** ▷ Variation in cases similar to variation in controls

      $k_{\min} \leftarrow k + 1$

  **else if** $\text{counter}_X \geq 1 \land \text{counter}_Y \geq 1$ **then** ▷ Scenario (iii) : NO variation in cases AND Controls

    $k_{\max} \leftarrow k - 1$

  **else** ▷ Scenario (iv) : NO variation in cases AND variation in controls

    **break**

**Output**: $k$

#### S1.2 Randomized PACA

The CCA step in PACA imposes the limitation $m \geq \max(n_0, n_1)$. In practice, there may be datasets in which this condition does not hold. In such cases, we can apply PACA on a subset of the samples, however, it is clearly sub-optimal to exclude data.

In order to address this limitation, we introduce randomized PACA (rPACA), a randomized variant of PACA, which can operate under the setup $m < \max(n_0, n_1)$ and learn case-specific variation for all $n_1$ samples in $X$. rPACA is based on the idea that the full sample covariance matrix of the cases (after adjustment for shared sources of variation) can be approximated by

**Algorithm S.A.2** Randomized PACA (rPACA)

**Input**: $X \in \mathbb{R}^{m \times n_1}$, $Y \in \mathbb{R}^{m \times n_0}$, $t$ number of iterations, $s < \min\{m, n_1, n_0\}$ subsample size, $q$ components of within-subsample case-specific variation , $k$ dimension of shared variation to remove.

$X'_{ij} \leftarrow X_{ij} - \frac{1}{m}\sum_{k=1}^{m} X_{kj},\ Y'_{ij} \leftarrow Y_{ij} - \frac{1}{m}\sum_{k=1}^{m} Y_{kj}$ ▷ Mean centering the matrices
$P \leftarrow \mathbf{0}$
**for** $n \leftarrow 1, ..., t$ **do**
  $X''_1, X''_2 \leftarrow \Pi_s(X')$ ▷ Randomly select $s$ train samples (1) and $n_1 - s$ test samples (2)
  $Y''_1, Y''_2 \leftarrow \Pi_s(Y')$ ▷ Randomly select $s$ train samples (1) and $n_0 - s$ test samples (2)
  **for** $r \leftarrow 1, ..., k$ **do** ▷ CCA

$$
\begin{aligned}
\hat{a}_r, \hat{b}_r \quad &= \quad \underset{a_r \in \mathbb{R}^B, b_r \in \mathbb{R}^B}{\operatorname{argmax}} (a_r)^\top (X''_1)^\top Y''_1 b_r \\
\text{s.t.} \quad & \quad \|X''_1 a_r\|_2 = 1, \|Y''_1 b_r\|_2 = 1 \\
& \quad \forall 1 \le h < r : X''_1 a_h \perp X''_1 a_r, Y''_1 b_h \perp Y''_1 b_r
\end{aligned}
$$

  $\hat{A} \leftarrow [\hat{a}_1, ..., \hat{a}_k]$
  $\hat{U}^0 \leftarrow X''_1 \hat{A}$
  $\tilde{X}_1 \leftarrow X''_1 - \hat{U}^0 (\hat{U}^0)^\top X''_1$
  $\hat{V}_{X_1}, \hat{Z}_{X_1} \leftarrow \text{PCA}(\tilde{X}_1{}^\top,\ \text{rank} = q)$ ▷ $q$ dims of within-subsample case-specific variation
  $\tilde{X}_2 \leftarrow X''_2 - \hat{U}^0 (\hat{U}^0)^\top X''_2$
  $\hat{V}_{X_2} \leftarrow \tilde{X}_2{}^\top \hat{Z}_{X_1}$
  $P' \leftarrow \begin{bmatrix} \hat{V}_{X_1} \\ \hat{V}_{X_2} \end{bmatrix}$ ▷ Stack projections to match order of $X$
  $P \leftarrow [P\ P']$ ▷ Hstack projection from each iteration
$C = PP^\top$
$\hat{V}_X \leftarrow \underset{||\mathbf{v}||_2 = 1}{\operatorname{argmax}} ||C\mathbf{v}||_2$ ▷ First eigenvector of the covariance matrix

**Output**: $\hat{V}_X$

learning projections from multiple subsets of the samples in the data. Finding the eigenvector that corresponds to the top eigenvalue of the estimated sample covariance matrix will then allow us to describe the most dominant variation in $X$ while accounting for the sources of variation that are shared with $Y$. Below, we provide a brief description of the rPACA algorithm; see full description in Algorithm S.A.2.

Briefly, rPACA begins by randomly splitting the data into two sets, each with a subset of the cases and a subset of the controls. rPACA uses one of the sets to learn the shared axes of variation between the cases and controls in the set, which are then used to adjust cases in both sets of the data. We then obtain the top $q$ (default: 10) PCs of both sets of cases and stack them into a single matrix; of note, we match the variances of the projections across the two subsets of cases, along each of the $q$ PCs, which alleviates possible variance shrinkage. Repeating this procedure multiple times using multiple splits of the data and combining the PCs across the multiple iterations allows us to use them as features, based on which we can approximate an (adjusted) sample-covariance matrix for the cases in $X$. This $n_1 \times n_1$ matrix then allows us to learn the top case-specific variation for all samples in $X$.

#### S1.3 Limitations

PACA has some limitations. First, because PACA automatically learns the directions of the shared sources of variation across $X$ and $Y$, it can, in principle, be robust to imbalances in confounders between $X$ and $Y$. However, like any unsupervised method, PACA is expected to be sensitive to scenarios in which there are severe confounder imbalances or in cases where confounders affect only $X$ and not $Y$. If both $X$ and $Y$ were collected as part of the same study and under proper randomization, such confounders, in principle, should not affect the data. Yet, in the event that such severe confounders are present in the data, they should be properly addressed if possible. For example, in the case of multi-site data, if certain sites collected only cases or only controls, then one should consider excluding the data from those sites prior to applying PACA. Otherwise, confounding effects due to site-specific variation may affect the ability of PACA to accurately capture the shared variation between cases and controls and the true phenotypic-specific variation. More generally, prepossessing is required when the confounder imbalance is severe. The approach of adjusting for confounding effects is context-specific, so users should employ field-specific best practices to select the appropriate prepossessing steps. This also includes the need to handle missing data, which is currently not internally addressed in our algorithm, but can, in principle, be addressed by developing a probabilistic extension of PACA. Finally, the robustness and calibration of PACA come at a cost: our assumption that the phenotype-specific signal is orthogonal to the confounders in the data may be violated in reality. We discuss this in the next subsection.

#### S1.4 Orthogonality assumption extended discussion

**Contrastive PCA is limited under the orthogonality assumption.** cPCA seeks to learn directions of variation that are enriched in a target dataset (e.g., cases) compared to a background dataset (e.g., controls), which reflects a contrastive learning paradigm by which a weak form of supervision (target versus background labels) informs the unsupervised learning of patterns in the target data. Using the same notations introduced earlier, given a parameter $\alpha > 0$, cPCA finds the top contrastive principal component (PC) by solving [2]:

$$v^* = \underset{v \in \mathbb{R}^m}{\operatorname{argmax}}\, v^\top X X^\top v - \alpha v^\top Y Y^\top v \qquad \text{s.t., } \|v\|_2 = 1 \tag{S.E.1}$$

In words, cPCA aims to find the dominant axis of variation in $X$ while controlling at a level associated with $\alpha$ for the variation this axis describes in $Y$. This framework allows us, in principle, to learn the case-specific variation $Z_X^1$ described in Eq. (1)-(2). Particularly, setting $\alpha = \infty$ allows cPCA to incorporate the orthogonality assumption and account for all shared sources of variation across $X$ and $Y$.

A solution to Eq. (S.E.1) while setting $\alpha = \infty$ requires that $\|Y^\top v^*\| = 0$ in order to avoid a negative infinity value for the objective. Therefore, $v^*$ must be orthogonal to the subspace spanned by the columns of $Y$, i.e., $v^* \in \text{null}(Y) := \{v | Y^\top v = 0_m\}$, where $0_m$ is an $m$-length vector of zeros. Effectively, since $X, Y$ are full-rank matrices due to the i.i.d. components of variation in Eq. (1)-(2), $v^*$ must be orthogonal to an $n_0$-dimensional subspace defined by to all samples in $Y$.

If $n_0 \geq m$ then $v^*$ must be orthogonal to the entire $m$-dimensional features space, hence we get $v^* = 0_m$ irrespective of the rank of $X$. Here, we are interested in the case where $\max(n_0, n_1) < m$, therefore $v^*$ is non-trivial, however, following the rank–nullity theorem, $v^*$ is restricted to an $m - n_0$ dimensional subspace of $\mathbb{R}^m$. Clearly, as $n_0/m$ approaches 1, cPCA will not allow us to capture the direction that spans the subphenotypic signals. Intuitively, this approach is suboptimal since it conditions on the entire $n_0$-dimensional subspace in $Y$ rather than on the lower-dimensional subspace that captures only the low-rank signals in the data. This suggests that a better approach is to condition only on sources of variation that exist in both $X, Y$, which are expected to be of lower dimension compared with the dimension of the observed data. This is exactly the key idea behind PACA.

**Orthogonality assumption limitations.** The orthogonality assumption may be violated if phenotypic heterogeneity is correlated with factors present in controls, such as population structure or other demographics. It can also occur due to study-specific artifacts, such as case/control misclassification (i.e., noisy/incorrect labeling of controls as cases or vice versa). In general, researchers should exercise caution when applying any contrastive learning approaches to low-confidence or noisy labeled data. As we show empirically, violation of the orthogonality assumption leads to a decrease in power to capture case-specific variation. However, we observe that PACA still outperforms alternative methods in scenarios of weak to moderate violation of the orthogonality assumption and is comparable in cases of severe violation (Fig. S3).

#### S2 PACA benchmarking

This section focuses on benchmarking PACA against other methods using both synthetic and real data. We begin by describing the simulation framework (S2.1) used to generate synthetic datasets for evaluating the performance of PACA and alternative methods. Next, we define the evaluation metrics (S2.2) employed to assess the methods' performance and explain how we implement the existing methods we compare PACA against (S2.3). We then introduce the datasets used for evaluation based on real data used in our benchmarking (S2.4). In the following subsections, we compare the power and Type 1 error calibration of PACA relative to existing methods and showcase PACA's ability to isolate known phenotype-specific heterogeneity (S2.5). Finally, we present results using existing methods for detecting asthma-specific heterogeneity in the GALA II data (S2.6).

##### S2.1 Simulation framework

We generate simulated control and case data with subtypes, as outlined here, for the power analysis in Subsection S2.5. For simulating datasets with two subtypes, we use the following model. Let $X_0 \in \mathbb{R}^{m \times n_0}, X_1 \in \mathbb{R}^{m \times n_1}$ be the data for controls and cases, respectively. To simulate this data, we assume the following model:

$$\begin{gathered} X_0 = WZ_0 + E_0 \\ X_1 = WZ_1 + W'Z_1' + E_1 \\ W \in \mathbb{R}^{m \times k}, \quad W' \in \mathbb{R}^{m \times 2} \\ Z_0 \in \mathbb{R}^{k \times n_0}, \quad Z_1 \in \mathbb{R}^{k \times n_1}, \quad Z_1' \in \mathbb{R}^{2 \times n_1} \\ E_0 \in \mathbb{R}^{m \times n_0}, \quad E_1 \in \mathbb{R}^{m \times n_1} \end{gathered} \tag{S.E.2}$$

We simulate the direction of the structure in the data, $W, W'$, as follows:

$$\begin{gathered} W'_{1,j'}, \ldots, W'_{m,j'} \sim \begin{cases} N(0, \sigma^2) & \text{with probability } \pi_0 \\ 0 & \text{with probability } (1 - \pi_0) \end{cases} \\ W_{1,j}, \ldots, W_{m,j}, \sim N(0, \sigma_j^2 \mathbf{I_m}) \\ \sigma_j^2 \sim \text{InverseGamma}(\alpha = 1.5, \beta = 1) \end{gathered} \tag{S.E.3}$$

where $\pi_0 \in [0, 1]$ defines the proportion of feature (signal density) that are active for each of the $j' \in [k_0]$ (here, $k_0 = 2$) latent dimensions, thereby controlling the density (sparsity) of the signal. In simulations, we generate data under the following signal densities: 0.01, 0.05, 0.25, and 0.5. $\sigma^2$ is set to 3 in all simulations. In the null case of no disease subtypes $W' = \mathbf{0}$. $\sigma_j^2$ defines the strength of the dense shares signal (most features are active, but at varying strengths) along each of the $j \in [k]$ shared latent components. In simulations, we set $k = 400$.

Next, we simulate the structure in the shared and unique structure. Let $k$ be the number of shared dimensions of structure across cases and controls. For $k' \leq k$, let $k'$ be the dim of the balanced shared variation, and $k - k'$ be the dim of the unbalanced shared variation. In the balanced axes of variation for $i \in \{1, \ldots, k'\}$:

$$\begin{aligned}(Z_0)_i, (Z_1)_i &\sim N(0, \sigma_i^2 \mathbf{I_n}) \\ \sigma_i^2 &\sim \text{InverseGamma}(\alpha = 1.5, \beta = 1)\end{aligned} \tag{S.E.4}$$

For the imbalanced axes of variation, when $k' \neq k$, $i \in \{k'+1, ..., k\}$:

$$\begin{aligned}(Z_0)_i &\sim N(0, \sigma_i^2 \mathbf{I_{n_0}}) \\ (Z_1)_i &\sim N(0, \tilde{\sigma}_i^2 \mathbf{I_{n_1}}) \\ \tilde{\sigma}_i^2, \sigma_i^2 &\sim \text{InverseGamma}(\alpha = 1.5, \beta = 1)\end{aligned} \tag{S.E.5}$$

We simulate the subtype structure, $Z_1'$, as a one-hot encoded matrix assignment of individuals to the subtypes. We alter the subtype signal strength by scaling the encoding's non-zero value to differing levels. In simulations, we generate data under the following signal strengths: 0.1, 0.25, and 1.

To explore scenarios where the case-specific subphenotypic signal is not orthogonal to the structure in the controls. To explore this, we can also add some of the case-specific structure to the controls $(\delta)^2 * W'Z_0'$. $W'$ is the same direction of structure as in the cases, $Z_0'$ encodes the random assignmnet of controls to one of the two subgroups, and $\delta \in [0, 1]$ controls the ratio of the variance of the subphenotypic signal in cases and the variance of the subphenotypic signal in controls. Taken together, we get the following model:

$$X_0 = WZ_0 + (\delta)^2 * W'Z_0' + E_0 \tag{S.E.6}$$

In simulations, we generate data under the following levels of orthogonality violation: 0, 0.25, 0.5 and 0.75.

Finally, we add normally distributed noise:

$$\begin{aligned}(E_0)_{ij} &\sim N(0, \sigma_{\epsilon 0}^2) \\ (E_1)_{ij} &\sim N(0, \sigma_{\epsilon 1}^2)\end{aligned} \tag{S.E.7}$$

where $\sigma_{\epsilon 0}^2, \sigma_{\epsilon 1}^2 \in [0, 1]$ controls the strength of the noise added to the controls and cases, respectively. In practice we set them to be equal to 0.1.

##### S2.2 Evaluation metrics

**Power.** For performance evaluation, we use Pearson's correlation between a putative subtype signal and the true subtype labels as a metric of power. To assess power, we use simulations, which allow us to control the presence/absence of imbalanced confounders and violation of the orthogonality assumption.

**Null Calibration.** We used a straightforward case/control label permutation approach to quantify the statistical significance of a putative subtype signal at a given $k$, or $\alpha$ for cPCA. For a fixed $k$, we build a null distribution of variances of the top PC by running PACA on all the permuted datasets. Then, we use this empirical null distribution to quantify the statistical significance of the observed variance of the top PC. This procedure should be able to reject the null when there is sufficiently strong variation unique to the cases. The exact same procedure is also used to quantify the statistical significance of the top PCA and cPCA components.

**Subtest.** We use Subtest [3], to check if a particular axis of variation defines an axis of genetically different architecture. Subtest is a statistical test to determine the existence of phenotypic subgroups in genetics. It does so by modeling the distribution of all SNP association statistics and fitting a mixture of Gaussians. Explicitly, it tries to fit SNPs to 3 Bivariate Gaussians: 1) a distribution of SNPs associated with neither the disease nor proposed subtype, 2) a distribution of SNPs only significantly associated with the disease and not the subtype, and 3) a distribution of SNPs which are either (i) associated only with the subtype or (ii) significantly associated with both the disease and proposed subtypes. Subtest fits a null model with a mixtures of models 1), 2) and 3)(i) and the alternative model with 1), 2) and 3)(ii). The difference of these model likelihoods provides a (pseudo) likelihood ratio (pLR). We can generate a null distribution by permuting the subtype values, generating the null test statistics, and fitting the Subtest to this data, which generates the null distribution for the pLR. This can be used to derive a p-value to quantify the significance the proposed subtypes.

Fundamentally, Subtest assumes that if valid subtypes exist, they should have some non-negligible proportion of SNPs that are associated with the subtypes and the primary disease. Subtest tries to find the existence of such subtypes. While we do not believe that this model encompasses the full space of all possible subtypes, this forms a well-defined approach for validating putative subtypes. In our application, we are using genetics to validate proposed subtypes uncovered by PACA in gene expression or genetic data. Therefore, we can use Subtest to test whether the top PACA component learns a valid axis of variation that defines a genetically differential subtype. Of note, Subtest also provides a way to test for subtype differences in each SNP using a Bayesian conditional False Discovery Rate (cFDR), by conditioning the subgroup test statistic on the primary phenotype test statistic for each SNP. We can use this procedure to select SNPs that tentatively discriminate between the putative subtypes.

##### S2.3 Evaluation of existing methods

We compared PACA to standard PCA, contrastiveVAE (cVAE) [4], and three variants of contrastive PCA (cPCA) [2]. For cVAE, we use the default architecture and hyper-parameters that were used by the authors [4], only modifying the input (and output) dimension size to match the feature size of the input data. We trained 10 cVAE models (different initialization), each for 50 epochs, and selected the best-performing run. When true subtype information was available, we considered the highest Pearson correlation. For datasets where subtype information was unknown, we used the Silhouette score as an internal metric (as described by Abid et al. [4]) to determine the best run.

For cPCA, the optimal choice of contrastive hyperparameter ($\alpha$) is unclear without prior subtype information. We, therefore, consider the best-case performance of cPCA, emulating a scenario where we have subtype information and can thus estimate the optimal contrastive value. To do this for every single run, we let cPCA automatically evaluate 10 different values of $\alpha$ and select the results yielding the highest correlation with the subtype signal. This version of cPCA, henceforth referred to as cPCA-opt, indicates cPCA with the optimal hyperparameter selection we identified, which reflects an overly optimistic benchmark of the expected performance of cPCA. Additionally, we tuned the contrastive hyperparameter following the procedure suggested by the authors [2]; henceforth, referred to as cPCA-auto. If we force cPCA-auto to return a single $\alpha$, it always, in our experiments, returns 0; as expected, this results in a non-contrastive, vanilla PCA. We, therefore, use the second suggested $\alpha$, which is the first non-zero value. Finally, we also assess the results of cPCA when setting the contrastive hyperparameter to the maximum ($\alpha = \infty$; referred to as cPCA-inf), which provides a view of cPCA that satisfies the orthogonality assumption we used in PACA; in practice, we set $\alpha = 10^6$.

##### S2.4 Datasets

We use genotype data from the UK BioBank (UKBB) [5] and gene expression data from the PsychENCODE ptoject [6, 7] to evaluate Type-1 error calibration in settings where no subtype signal exists, allowing us to assess the methods' performance under null conditions. To estimate power (positive-control validation analysis), we use the aforementioned datasets, as well as the genetic data in PsychENCODE and DNAm data from the Rahmani et al. [8] and Hannum et al. [9] studies (described below). We also briefly use the HapMap dataset to illustrate miscalibration.

**HapMap.** We used genotypes from the HapMap project (release 23a, filtered). A total of 120 samples (60 YRI and 60 CEU founders) and 1,872,641 SNPs were available for the analysis.

**UK BioBank.** We used a subset of the UK BioBank genotypes dataset with 291,273 unrelated white British individuals and 459,792 SNPs, filtered and processed as described elsewhere [10]. We used this full dataset to calculate the first 10 PCs that were used as covariates in all genome-wide association (GWAS) analyses needed for our application of Subtest. We used Field 20002 (Non-cancer illness, self-reported; instance 1) in the UK BioBank phenotype data to obtain disease type and status. We classify individuals as (non-disease) controls if they have no reported diseases in Field 20002, from which we randomly select 5,000 individuals that we use as controls for all associated analyses. We also use Field 20002 to find 2,500 mutually exclusive cases of coronary artery disease (CAD) and rheumatoid arthritis (RA) for each (randomly sampled when total cases >2,500). Note that we define CAD as any individual who reported angina or myocardial infarction. We created a 10,000 SNPs Panel with 2,000 RA, 2,000 CAD, and 6,000 Null SNPs. We selected the top 2,000 mutually exclusive SNPs for each disease from the Neale Lab GWAS summary statistics (v3) [11], by p-value. The selection was based on cross-referencing against a linkage disequilibrium (LD) clumped list, using a $\pm$ 100 kilobase window and an LD threshold of $< 0.1$. The SNPs summary statistics were ranked by their p-values, and only the SNPs present in our dataset were considered. For the Null SNPs, we randomly chose 6,000 SNPs from an LD-pruned (LD$< 0.1$) list that had no overlap with the top (13,809 Bonferroni passing) disease SNPs.

**PsychENCODE.** The PsychENCODE data we used contains genotype, gene expression, and covariate information for 644 control, 472 schizophrenia (SCZ), 172 bipolar disorder (BP), and 33 Alzheimer's disease (AD) samples. We only used SCZ, BP, and data from control samples. We used the raw count gene expression levels of all 25,774 genes available in the data as the input to the benchmarked methods, and we used a set of 217,863 LD pruned ($< 0.1$) SNPs for the Subtest analysis.

**DNAm datasets.** We used two HumanMethylation450 BeadChip array-based datasets. A dataset of Latino samples from the GALA II cohort (a subset of which overlaps with the EPIC GALA II samples used in our main asthma study; n=573; GEO accession ID GSE77716) [8], and a dataset from the Hannum et al. study with European population (n=656; GEO accession ID GSE40279) [9]. We considered the top 50,000 most variable CpGs in the data for evaluation. Prior to analysis, we excluded outlier samples that demonstrated extreme values in the top two PCs of the data (values in PC1 or PC2 exceeding 3 standard errors), which resulted in n=558 and n=651 samples for the GALA II and Hannum datasets, respectively. The GALA II dataset includes 77 samples labeled as other or mixed Latino; these were excluded from the final evaluation, in which we only considered Mexican and Puerto Rican samples.

##### S2.5 PACA is calibrated and well-powered for identifying disease subtypes

**Assessment of Type I Error Rates.** We first evaluate calibration in null scenarios of no subtypes (i.e., no case-specific variation). A desired property of a method for capturing phenotypic heterogeneity is being calibrated under the null. That is, given data with no phenotypic heterogeneity (i.e., the null case) and a prespecified confidence level, the method should be able to determine that no phenotypic heterogeneity exists while controlling for the target type I error rate that corresponds to the prespecified confidence level. In order to evaluate calibration, we applied PACA to data with no phenotypic heterogeneity by randomly sampling two groups of healthy individuals we labeled as "cases" and "controls" from each of the UK Biobank and the PsychENCODE datasets. Unlike the alternative methods, PACA was always calibrated and maintained target Type I error rates (Fig. S1).

**Assessment of power.** To benchmark the power difference across methods, we generated synthetic case-control data with case-specific heterogeneity. We first generated synthetic case-control data, in which cases are assigned into two subtypes. Each dataset combined two types of variation: (i) multiple components of variation common to cases and controls (shared variation) and (ii) case-specific variation that differentiates between the two subtypes of cases with varying signal strengths and sparsities (i.e., the fraction of features in the data defining the subtypes). For each generated dataset, we evaluated the absolute Pearson correlation between the binary classification of cases into subtypes and the first component of each method.

We found that PACA substantially outperformed the alternative methods under all combinations of the strength and sparsity of the subtype signal in the data, except for cases where the subtype information is strong and defined by a large fraction (50%) of the features in the data, in which case it was on part with the best performing methods (Fig. S2). PACA provides the most notable improvement over the competing methods in scenarios where the two subtypes are imbalanced in prevalence (Fig.S2). Similarly, the simulation results show that rPACA outperforms alternative methods under most conditions but has weaker performance than PACA, likely due to shared dimension estimation errors in the smaller subsets of data.

We further benchmarked the different methods under violations of the orthogonality assumption, that is, in scenarios where the subtype signal is not case-specific but is rather correlated with the shared variation and affects control samples as well. While we observed an expected substantial performance decrease for all methods in that case (Fig. S3), compared to other methods, PACA demonstrated significantly better performance under weak to moderate correlation between the subtype classification and the shared variation (Fig. S3b). Severe violation of the orthogonality assumption, however, rendered all methods ineffective in identifying the underlying subtype information (Fig. S3c,d).

Finally, in order to gain more insight into the superior performance of PACA, we compared the simulated dimension of shared variation between cases and controls ($k_0$) to the dimension of shared variation that was removed by PACA. We found that in cases of weak phenotypic heterogeneity signals, PACA tends to adjust the data for a number of axes of shared variation that approximately matches the true simulated dimension of shared variation (Fig. S23). As the strength of the signal increases, a larger subspace of the space that defines the shared sources of variation is expected to include signals that are weaker than the phenotypic heterogeneity signal. PACA leverages this insight and estimates the "effective" (i.e., minimal) dimension of shared variation that needs to be removed in order to detect the phenotypic heterogeneity signal (i.e., rather than necessarily removing all the shared variation; Algorithm S.A.1); indeed, our analysis confirms that PACA adjusts for fewer axes of shared variation as the signal strength and density increase (Fig. S23).

**Recapitulating population structure from DNAm.** The most dominant source of variation in DNAm from heterogeneous tissues such as blood is known to be cell-type composition [8]. However, it has been previously reported that a large number of methylation CpGs are strongly correlated with population structure [12, 13]. We, therefore, tested if PACA is powered to capture population stratification from heterogeneous whole-blood DNAm samples that were collected from individuals of Mexican and Puerto Rican ethnicities in GALA II. To that end, we applied PACA and other methods to this target dataset, contrasting it with a background dataset collected from European individuals in the Hannum et al. dataset. In this scenario, the two different populations in the GALA II dataset, which do not exist in the Hannum data, emulate "subtypes".

PACA achieved the best performance in capturing the population stratification in the data (r=0.44; P=1.2e-24) (Fig. S4a). Conversely, even the best performing cPCA variant in this case, cPCA-inf (which permits no background variation), failed to capture the target variation (r=-6.3e-2, P=0.17)(Fig. S4a), underscoring the need to account for an adequate dimension of shared variation between the target and background data. Similarly, cVAE was also unable to capture meaningful population stratification, instead collapsing the latent representation collapses towards zero (Fig. S4a).

**Uncovering genomic heterogeneity in gene expression data.** We broaden our analysis of PACA's performance by attempting to delineate between schizophrenia and bipolar disorder. The substantial shared polygenic basis of these disorders [14] makes their stratifying from a mixed sample of cases a notably difficult task. We applied PACA to a mixture of gene expression profiles from the PsychENCODE data [6, 7]. We pooled together schizophrenia (n=472) and bipolar (n=172) samples as the "cases" group to model a synthetic disorder with two subtypes. For the background group, we included all controls in the data (n=644).

We confirmed that the orthogonality assumption is violated in this case: the synthetic disorder subtype was correlated with the RNA sequencing technique (library preparation) for the samples (poly(A) enrichment or ribosomal RNA depletion; r=0.12, P=0.003). A PCA analysis reveals that the first two PCs of the case group perfectly separate the samples by the library preparation type (Fig. S24). In particular, PC1 is strongly correlated with the library preparation (r=0.70, P=2.2e-16). As a result, PC1 is also correlated with the synthetic disorder subtype (r=-0.26, P=1.5e-11) due to the violation of the orthogonality assumption. For that reason, we tested whether PACA and the other methods can capture the subtype signal beyond the part that is correlated with the library preparation by adjusting for it in a linear regression analysis.

The first contrastive PC of cPCA-opt (the best-performing cPCA version) did not capture a substantial subtype signal (P=0.056; linear regression) for the $\alpha$ resulting in the highest correlation with the subtype (Supplementary Tables S17 and S18). We also confirmed that cVAE yielded no significant association with the subtype status after correcting for library preparation. On the other hand, the first PACA PC did successfully identify the subtype signal while accounting for the library preparation confounder (P=2.6e-07; linear regression; Fig.S4b). Using Subtest [3], we further found that PACA PC1 isolated heterogeneity in the genetic architecture (P=1.4e-6) of the bipolar and schizophrenia cases, despite their extensively shared polygenic architecture [14]. Overall, these findings underscore PACA's power and effectiveness in identifying condition-specific heterogeneity, even under violation of the orthogonality assumption.

**Capturing phenotypic heterogeneity in genetic data.** Common genetic variation and specifically SNPs are notorious for their small effect sizes in complex disease [15, 16]. This makes the task of finding subtypes solely based on genetic data particularly hard. We, therefore, evaluated the potential utility of PACA in identifying phenotypic heterogeneity from SNPs. To that end, we mixed genotype arrays of CAD cases (n=2,500) and RA cases (n=2,500), which we pooled together from the UK Biobank data [5] into one group. Due to the large number of genetic variants in typical genotype data, we assumed that prior work had identified sets of variants likely associated with phenotypic subtypes. Therefore, our analysis used a CAD/RA-enriched panel of SNPs as features.

We applied PACA to this mixed group of cases while contrasting it with a group of healthy control individuals from the UK Biobank (n=5,000), and we found that the top PACA component is enriched for correlation with CAD- and RA-associated SNPs that represent a sub-phenotypic genetic heterogeneity in this case (Fig. S25**a**). Finally, we applied Subtest to the data, which further confirmed that the top PACA component defines a spectrum of phenotypic heterogeneity that presents differential genetic architecture between the CAD and RA cases (P <4.201e-40). Furthermore, we identified 211 SNPs (Subtest conditionalFDR < 0.05; Fig. S25) likely contributing to the difference in the genetic basis of these diseases along the top

PACA component, of which 107 are *a priori* RA-associated SNPs and 39 are CAD-associated SNPs.

##### S2.6 Applying alternative contrastive methods for asthma patient stratification

We juxtapose our asthma methylation score results in GALA II against the top 2 PCs from PCA (case only; served as a naive baseline), cPCA-inf, cPCA-auto, and cVAE (Tables S3 and S5). cPCA-opt cannot be used in this analysis since we wish to identify *de-novo* heterogeneity, and therefore, we do not have a reference variable for evaluating the performance of cPCA under different regularization hyperparameters.

We found that the top PC of PCA is primarily correlated with neutrophil counts (r=-0.89, p=3.9e-43), lymphocyte counts (r=0.86, p=3.9e-37), and BMI (r=-0.23, p=1.5e-08), while the second PC is strongly correlated with BEC (r=0.85, p=2.8e-35) and Nitros oxide pollution levels (r=0.59, p=1.8e-11). The second PC is also correlated with BDR (r=0.23, p= 6.2e-9), however, this association disappeared (p=0.28; linear regression) after adjusting for BEC. These results indicate that the top two PCs of the data capture, as expected, cell-type composition.

Similarly, cPCA-auto cPC1 was significantly associated with neutrophil counts (r=0.89, p=1.3e-43), lymphocyte counts (r=-0.86, p=3.2e-37), and white blood cell count (r=0.42, p=1.0e-6), and cPC2 was associated with BEC (r=-0.86, p=1.1e-36), Nitros oxide pollution levels (r=0.-60, p=9.9e-12), and BDR (r=-0.23, p=7.50E-09). As with PCA, the association with BDR is no longer significant (p>0.17; linear regression) when adjusting for BEC, indicating that cPCA-auto captures cell-type composition as well. Neither of the top two components of cPCA-inf was significantly correlated with any of the clinical phenotypes after correcting for multiple hypotheses (Bonferroni).

Finally, we observe that the variation in the top component of the cVAE model is primarily correlated with by sex (r=-0.33, p=1.0e-16); the second component is correlated with sex (r=0.42, p=4.2e-27), BEC (r=-0.44, p=4.2e-7), baseline FEV1/FVC ratio (r=-0.27, p=6.0e-12), and educational attainment of father (r=0.17, p=2.3e-5). Although sex, BEC, and baseline FEV1/FVC ratio are predictive of asthma outcomes [17], these components are not significantly correlated with BDR after correcting for multiple hypotheses.

### S3 GALA II and SAGE II extended details

#### S3.1 Extended DNAm preprocessing protocol

Bad-quality methylation data points were defined as those with a detection p-value $> 1 \times 10^{-6}$ or a number of beads less than 3. Low-quality CpG probes were defined as those having $\geq 5\%$ of bad-quality data points across samples. Low-quality samples were defined by one or more of the following criteria: 1) $\geq 5\%$ of bad-quality data points across CpGs, 2) total bisulfite intensity less than 3 standard deviations below the bisulfite controls, or 3) outliers in bisulfite intensity or beta-value distribution. These low-quality data points, probes, and samples were removed.

Further quality control and data processing were conducted using the `ENmix` R package (version 1.22.0) [18] as follows. Background correction was performed using the out-of-band (oob) method, and intensities were normalized to remove technical inter-array variation via quantile normalization. Dye and probe-type biases were corrected using the Regression on Correlated Probes (RCP) method and the Regression on Logarithm of Internal Control (RELIC) method, respectively. Methylation intensities were used to compute beta values (ranging from 0 to 1). Outlier methylation data points for each CpG (identified by the 3×IQR rule) were set as missing values. CpGs and samples with a missingness rate $> 5\%$ and $> 10\%$, respectively, were removed, and the remaining missing values were imputed using the k-nearest neighbor method.

Related individuals were identified based on WGS data. These, as well as individuals with discordant sex between the reported and methylation-predicted sex, were removed. Using the ewastools R package (version 1.7) [19], individuals with a mixed distribution of beta values for genotyped SNPs in EPIC (59 SNPs) were removed due to potential cross-sample contamination. Probes located on sex chromosomes (X and Y) and those discarded from the Illumina EPIC manifest v1.0 B4 file were removed. Potentially problematic probes that may capture artifacts other than methylation were flagged. These include probes with a multimodal distribution of beta values, cross-reactive or non-specific probes, and those potentially affected by genetic variation. Potentially polymorphic probes were defined as those containing a SNP: 1) at the CpG site, 2) at a single base extension (SBE) from the CpG site, or 3) within the probe with a minor allele frequency (MAF) $> 1\%$, based on the Illumina EPIC manifest file v1.0 B4. MAFs were estimated for each cohort separately. These potentially problematic probes were filtered out before analyses.

#### S3.2 Hospitalizations and ER visits

Hospitalization and emergency room (ER) visit data were directly obtained through interviewer-assisted questionnaires. Hospitalizations were determined by asking, ”In the last 12 months, has [CHILD] been hospitalized because of asthma?” (0 = no, 1 = yes), while ER visits were assessed with the question, ”In the last 12 months, has [CHILD] received any asthma care in an Emergency Room, a physician’s office, or clinic that was not scheduled at least 24 hours ahead of time?” (0 = no, 1 = yes).

##### S3.3 Exacerbations

We created a composite asthma exacerbation score based on the definition set by the American Thoracic Society and European Respiratory Society [20]. Briefly, we assigned points for reported asthma-associated hospitalizations in the last 12 months (one point for one hospitalization, two points for 2-4 hospitalizations, and 3 points for 5 or more hospitalizations), asthma-associated emergency department, physician's office, or clinic visit that was not scheduled at least 24 hours ahead of time in the last 12 months (one point for one visit, two points for 2-4 visits, and 3 points for 5 or more visits), and past-year oral steroid use (one point was given if the participant used oral steroids in the past 12 months). To avoid double-counting for oral steroid prescriptions, a point was given to those who used any oral steroid in the last 12 months only among participants who did not report a history of emergency visits or hospitalizations for asthma. Points were then summed to derive a composite score of values ranging from 0-6.

##### S3.4 BMI, ICS, and ancestry fractions

The measurements of weight and height were used to calculate BMI percentiles using sex- and age-specific curves for each participant at the study visit [21]. We consider ICS to be true if the subject has taken any inhaled controller medication, excluding Advair, in the past 12 months. Global ancestry proportions were estimated from whole-genome sequencing data using the protocol described in the online supplement by Mak et al. (2018) [22].

#### S4 Alternative models for predicting bronchodilator response

We evaluated several alternative predictive models for BDR. A logistic regression model using cell-type composition, a significant source of variation in DNAm data [23], underperformed compared to the two-biomarker models tailored for patients with high DNAm scores (Supplementary Table S6). A DNAm-based regularized logistic regression model using the same 7,662 CpGs that define our DNAm score outperformed the two-biomarker models in predicting BDR in GALA II (ROC AUC 0.83). However, unlike the simple two-biomarker model for patients with high DNAm scores, the performance of this model did not replicate in the SAGE II cohort (ROC AUC 0.65), presumably due to overfitting to confounding effects. These results further underscore PACA's effectiveness in eliminating unknown confounders, by using contrastive learning to remove sources of variation shared across cases and controls. Interestingly, both the cell-type composition and DNAm-based regularized logistic regression models performed better when evaluated in patient groups with high DNAm scores, further underscoring the robustness of the proposed DNAm score in stratifying clinical outcomes (Supplementary Table S6).

### S5 Computational deconvolution of the seven plasma cell-related genes extended details

We applied CIBERSORTx (`GEP` mode) [24] to the transcripts per million (TPM) normalized raw counts of the whole-blood RNA sequencing data in GALA II and SAGE II and to the quantile-normalized levels of the microarray data in the EXTRA data. We used the immune cell reference panel LM22 [25] to estimate cell-type expression levels and proportions of 12 immune cellular compartments: neutrophils, eosinophils, monocytes, macrophages, dendritic, CD4+ T, CD8+ T, regulatory T, B, plasma, natural killer, and mast cells. We provide the p-values for the cell-type-level expression of the genes TNFRSF13B, TNFRSF17, IGHV6-1, IGLV5-45, ITM2C, TXNDC5, and TXNDC11 in Supplementary Table S16.

Next, we calculated a composite T2ABC score we defined as the total expression of the seven T2ABC genes above, which we confirmed to be predominantly expressed in plasma cells (Supplementary Fig. S13 and S14; Supplementary Table S16). To confirm an association – between the seven-gene score based on whole-blood expression and the DNAm score – that is statistically driven by plasma cell-level expression, we applied TCA (version 1.2.1) [26]. Since TCA assumes the data is normally distributed, we used per-gene log1p-transformed expression levels for the composite score, which we eventually scaled to have variance 1.

We first applied the `tca` function to learn a deconvolution model for the seven-gene T2ABC score, using the cell-type proportion estimates from CIBERSORTx as an input. We accounted for age, sex, BMI, education level, African and European fractions based on genetic ancestry, Mexican ethnicity (only for GALA II), ICS use, and OCS use. Then, to test the statistical effect of the seven-gene T2ABC score on the DNAm score, we applied the `tcareg` function with the T2ABC score as explanatory variable for the DNAm score, which served as the outcome variable. We accounted for the same set of covariates described above, as well as the estimated cell-type proportions, while using the default parameters of `tcareg`, except that `fast_mode` was set to `FALSE`. (We opted for the slower, yet more accurate option since we tested a single gene.) Lastly, since we expected elevated T2ABC scores to correspond to elevated DNAm scores, we considered a one-sided alternative hypothesis; in practice, this was implemented by dividing the p-values reutrned by `tcareg` by 2 (after confirming the expected direction of effect). This analysis confirmed that plasma-cell expression of the seven-gene T2ABC score is associated with the DNAm score in both GALA II (P=0.038) and SAGE II (P=7.5e-4).

#### Part II

### Supplementary Figures

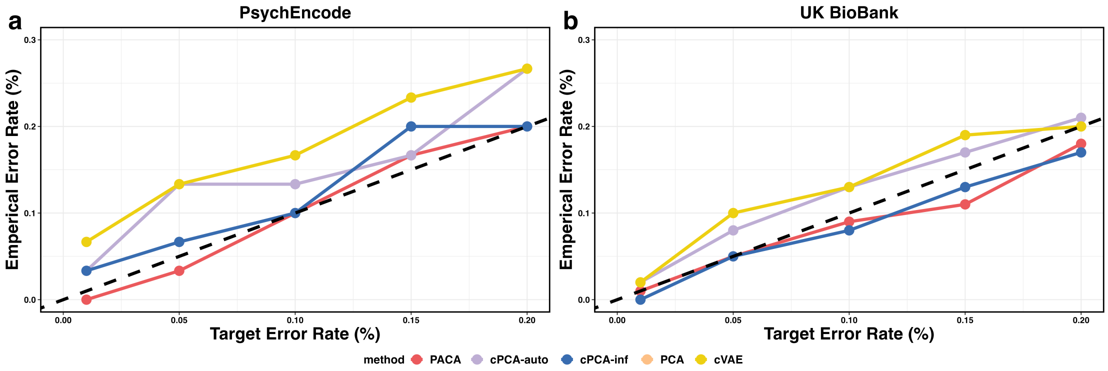

**Supplementary Figure S1:** Evaluation of type I error calibration under the null case of no subphenotypic variation. **(a)** The type I error calibration of PACA, cPCA with $\alpha = \infty$ (cPCA-inf), cPCA-auto, cVAE, and PCA based on genotype data from non-diseased unrelated White British individuals from the UKBB (N=2,000 and a randomly selected subset of 10,000 SNPs) over 100 simulations. **(b)** Type I error rates based on gene expression data on control samples from PsychENCODE (N=1,000 and 25,774 genes) over 30 simulations. The x-axis represents the target type I error, and the y-axis corresponds to the observed type I error at the target error rate. Dashed lines represent type I calibration.

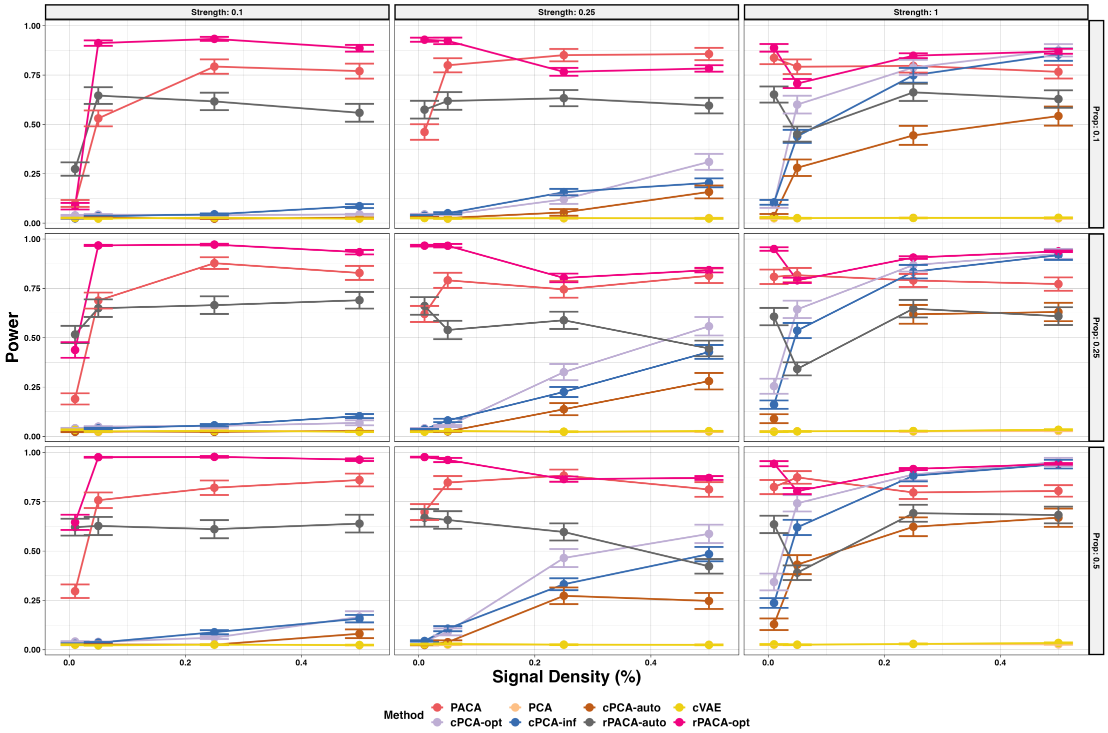

**Supplementary Figure S2:** Evaluation of statistical power in capturing subphenotypic signal under simulations in the presence of two subtypes ($n = 2000, m = 2000, k = 400$). Performance was evaluated for PCA, cVAE, PACA with automatic $k$ selection, rPACA with fixed optimal $k = 400$ (rPACA-opt), rPACA with automatic $k$ selection (rPACA-auto), cPCA with the best performing regularization parameter $\alpha$ among a range of 10 values suggested by cPCA (cPCA-opt), cPCA while setting the regularization parameter to $\alpha = \infty$ (cPCA-inf), and cPCA with the first non-zero suggested alpha (cPCA-auto). Presented is the linear correlation of the top component (power) of each method with the simulated subtype as a function of the signal density (lower values correspond to more sparse signal), across a range of signal strengths (columns; low=0.1, medium=0.25, high=1.0) and levels of imbalance (Prop) between the prevalence of the two subtypes (rows). The performance of every combination of parameters was averaged across 100 simulated datasets; vertical bars represent standard errors.

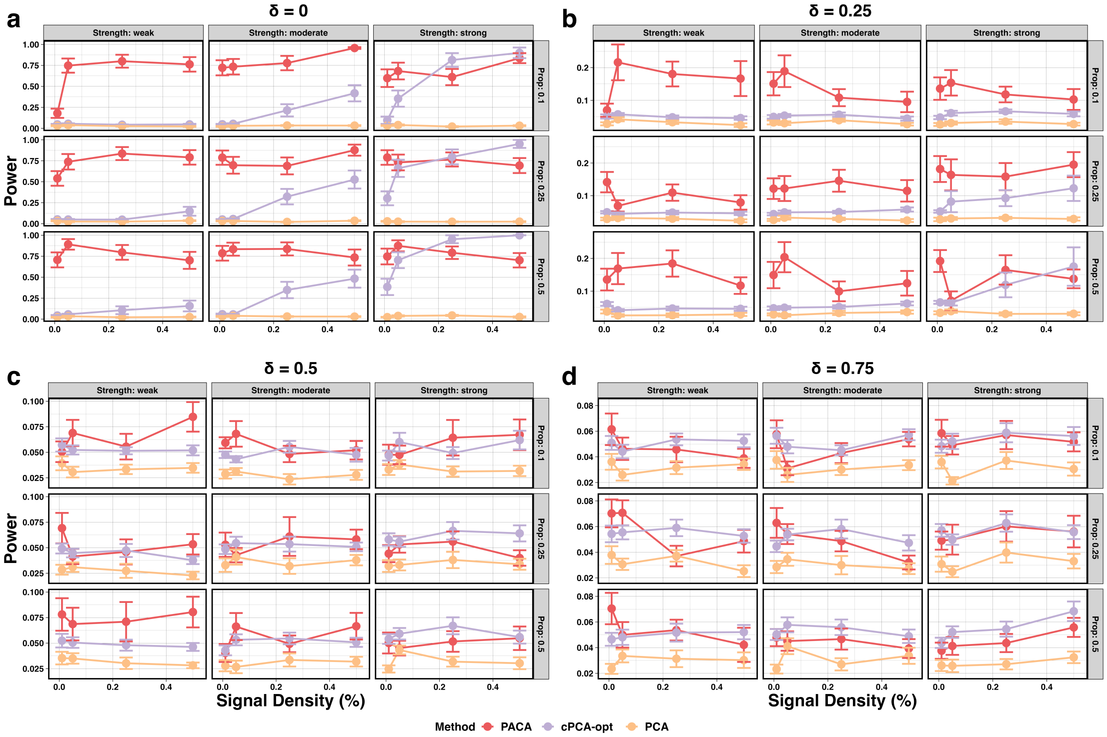

**Supplementary Figure S3:** Evaluation of statistical power in capturing subphenotypic signal under the scenario where the assumption of orthogonality between the case-specific variability and the case-control shared variability is violated. For each of several levels of violation of the orthogonality assumption (denoted by $\delta$; higher levels indicate more significant violation), presented is the correlation of the different methods with the subtype signal as a function of the signal strength, case subtype proportion (Prop), and signal density (lower values correspond to more sparse signal). The results are based on simulated data with $N = 1,400$, $k_0 = 300$, and 2,000 features, averaged over 20 simulations. The columns are evaluations at a fixed signal strength, the rows are evaluations at fixed case proportions, each x-axis indicates the corresponding signal density, and the vertical bars represent standard errors. For visualization purposes, only the top-performing methods were included.

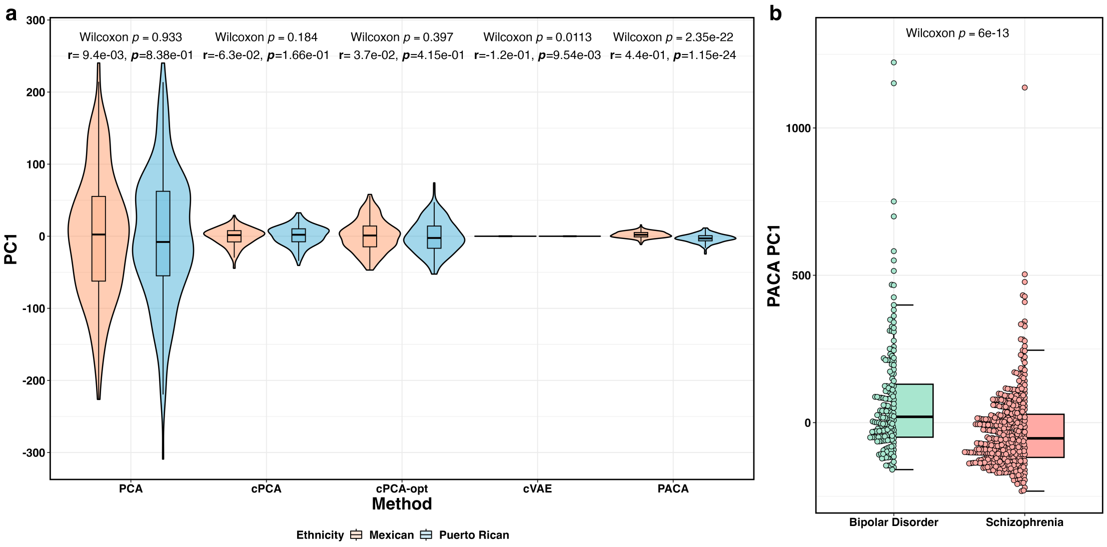

**Supplementary Figure S4:** **(a)** Capturing population heterogeneity (emulating subphenotypic variation) in whole-blood DNAm data from admixed individuals as "cases" and European individuals as "controls". Presented are the first two components found by standard PCA, PACA, which accounted for an automatically estimated 16-dimensional signal of shared variation, cPCA with the best performing regularization level among a range of values suggested by cPCA, and cPCA with the regularization level set to $\alpha = \infty$. The colors represent the self-identified ancestry. **(b)** Stratifying bipolar and schizophrenia cases from gene expression in the PsychENCODE data. Boxplots overlaid with swarm plots show the distribution of the PACA PC1 values among bipolar disorder (green) and schizophrenia (red).

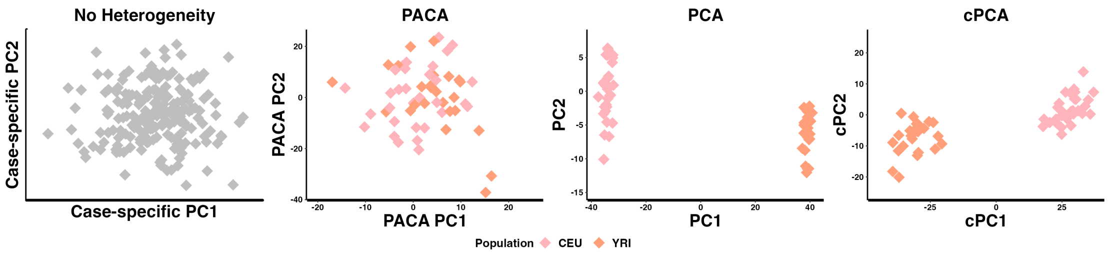

**Supplementary Figure S5:** Evaluation of the top two components of PCA, cPCA, and PACA under a scenario with no phenotypic heterogeneity, in which genotype samples from the HapMap data were randomly assigned with a "case"/"control" status (58 YRI and 58 CEU individuals; based on a randomly selected set of 10,000 SNPs). The left-most plot with gray points illustrates the expected output from a calibrated method for detecting phenotypic heterogeneity. Two outlier samples from HapMap results were excluded for visualization purposes.

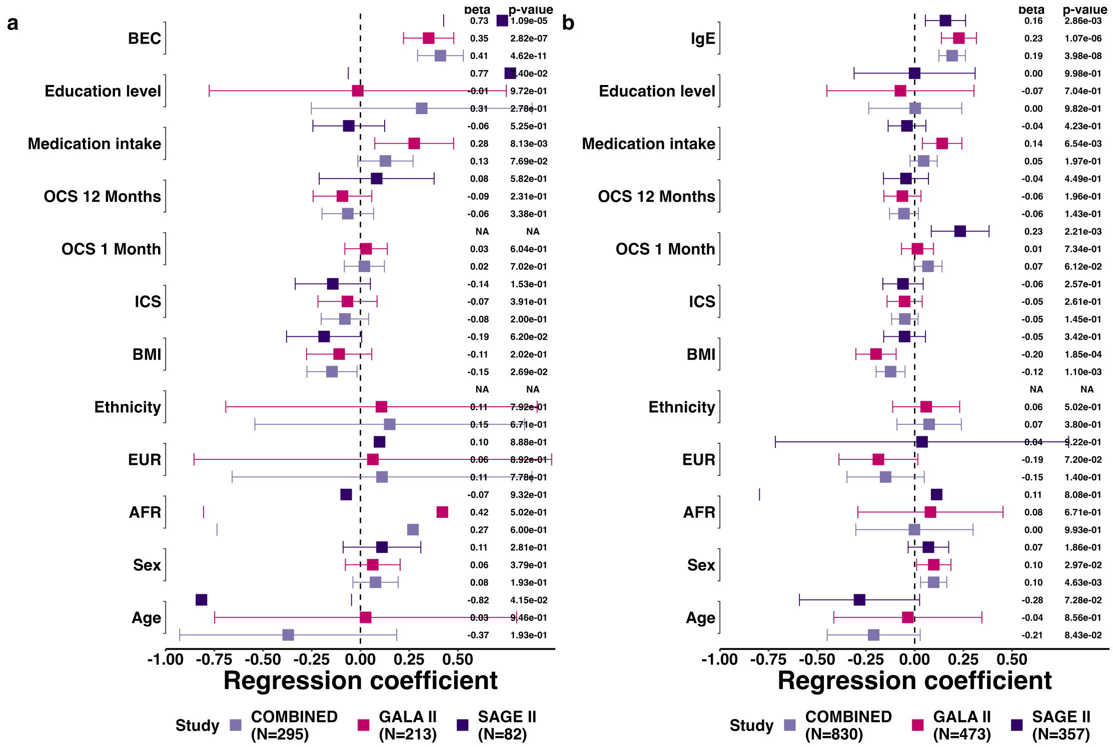

**Supplementary Figure S6:** Linear regression coefficients and 95% confidence intervals for models with the DNAm score as the outcome and **(a)** BEC or **(b)** log total serum IgE as the variable of interest. Models are adjusted for demographic and clinical variables, and results are stratified by study (GALA II, SAGE II) or combined (COMBINED).

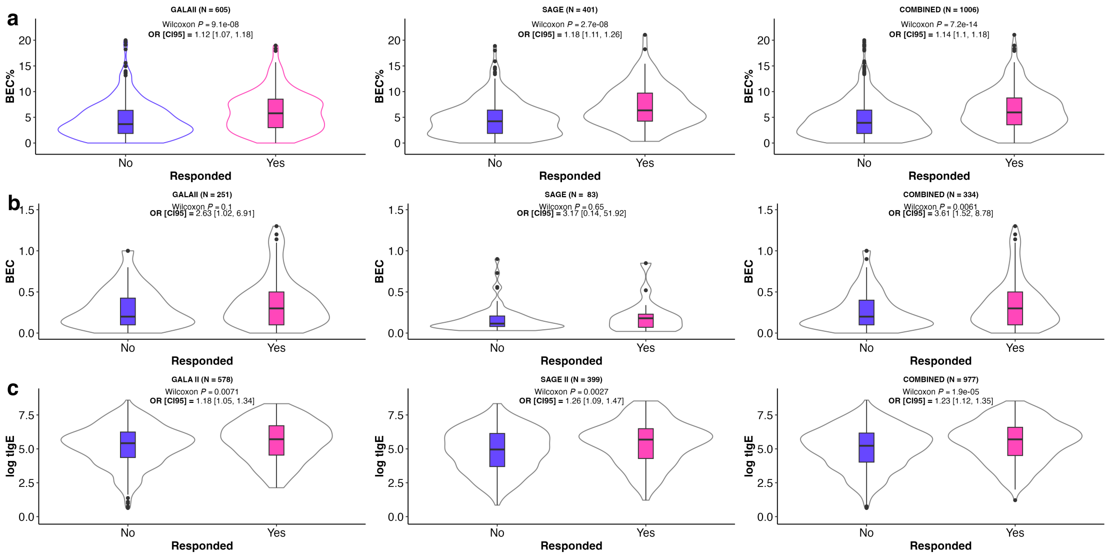

**Supplementary Figure S7:** Evaluation of the predictive value of BEC and IgE for BDR. Violin plots show the distributions of **(a)** imputed BEC (%), **(b)** BEC (cells/$\mu$L; measured for a subset of the samples), and **(c)** log-transformed total serum IgE levels, stratified by BDR response (BDR $\geq$ 12%). Results are also stratified (columns) by study (GALA II, SAGE II) or combined (COMBINED).

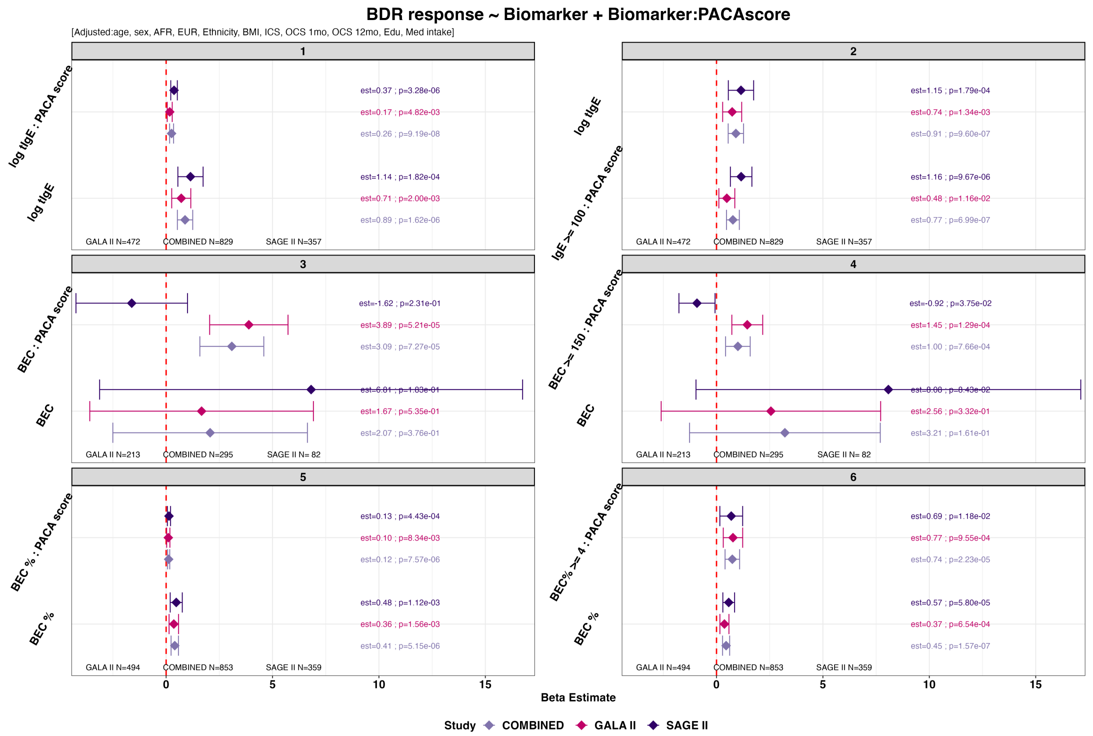

**Supplementary Figure S8:** Modeling BDR as a function of statistical interactions between the DNAm score and different biomarker definitions based on BEC and IgE. Each panel displays results from a linear regression model incorporating both a linear effect and an interaction term, using a different biomarker definition in each case. Presented as estimated effect sizes with 95% confidence intervals, results are stratified by study (GALA II, SAGE II) or combined (COMBINED. All models were adjusted for demographic and clinical covariates.

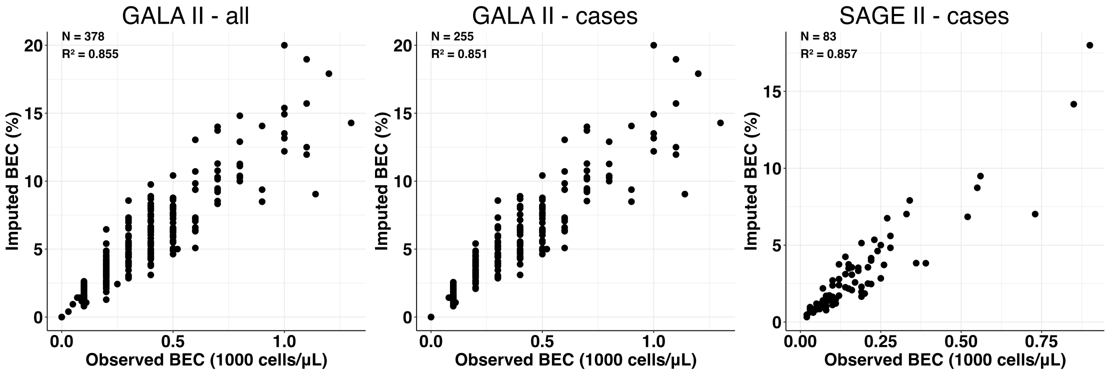

**Supplementary Figure S9:** Comparison of measured BEC (cell counts) with imputed BEC proportions (BEC%) across different subsets of the **(a)-(b)** GALA II (all samples and cases only) and **(c)** SAGE II cases, where complete blood cell counts including BEC were measured.

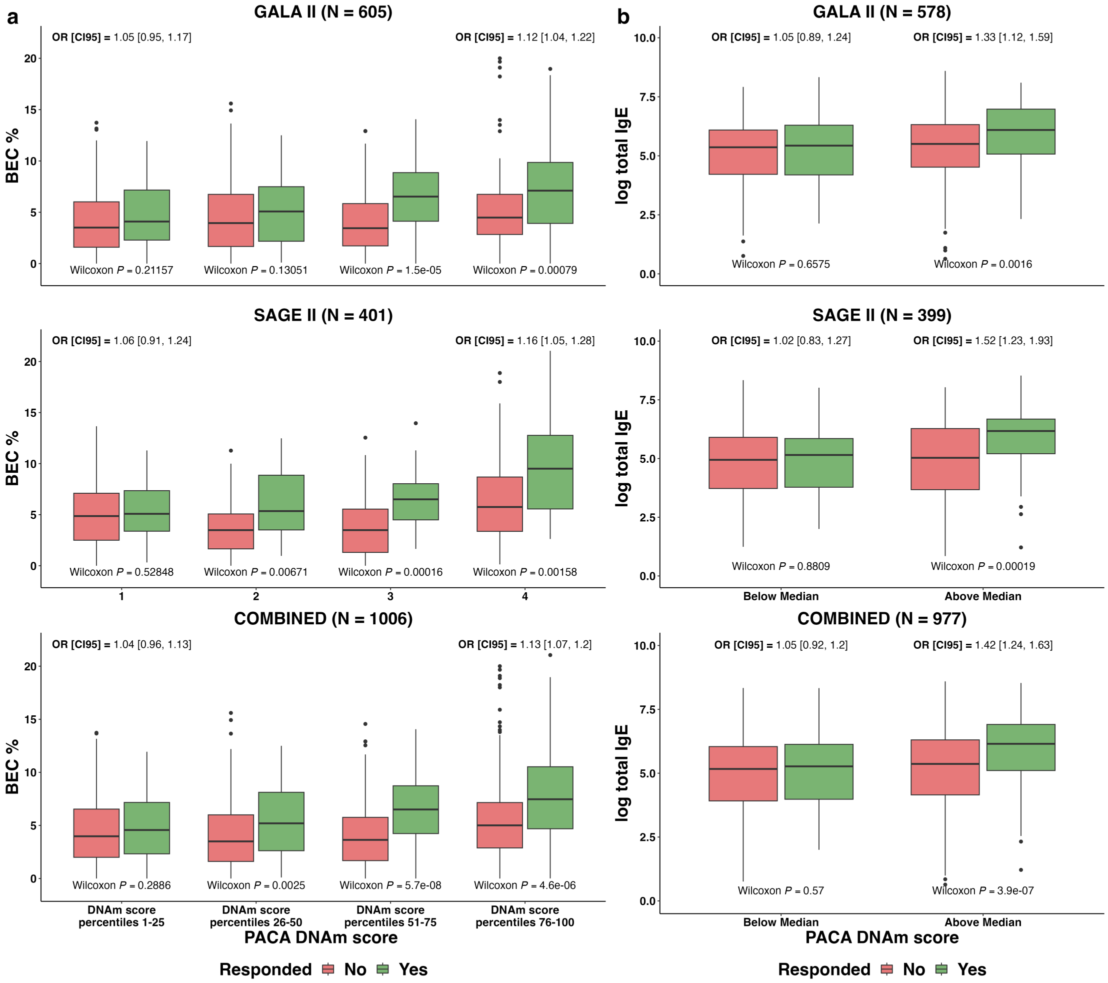

**Supplementary Figure S10:** Differential predictive value for BEC and IgE in predicting BDR along the DNAm score spectrum. Grouped boxplots compare the distributions of **(a)** imputed BEC proportions (BEC%) across quartiles of the DNAm score and **(b)** (log) IgE levels across below- and above-median DNAm scores, stratified by BDR response (BDR $\geq$ 12%). Low Wilcoxon rank-sum p-values and high odds ratios indicate the biomarker is predictive of BDR for patients in the respective percentile range.

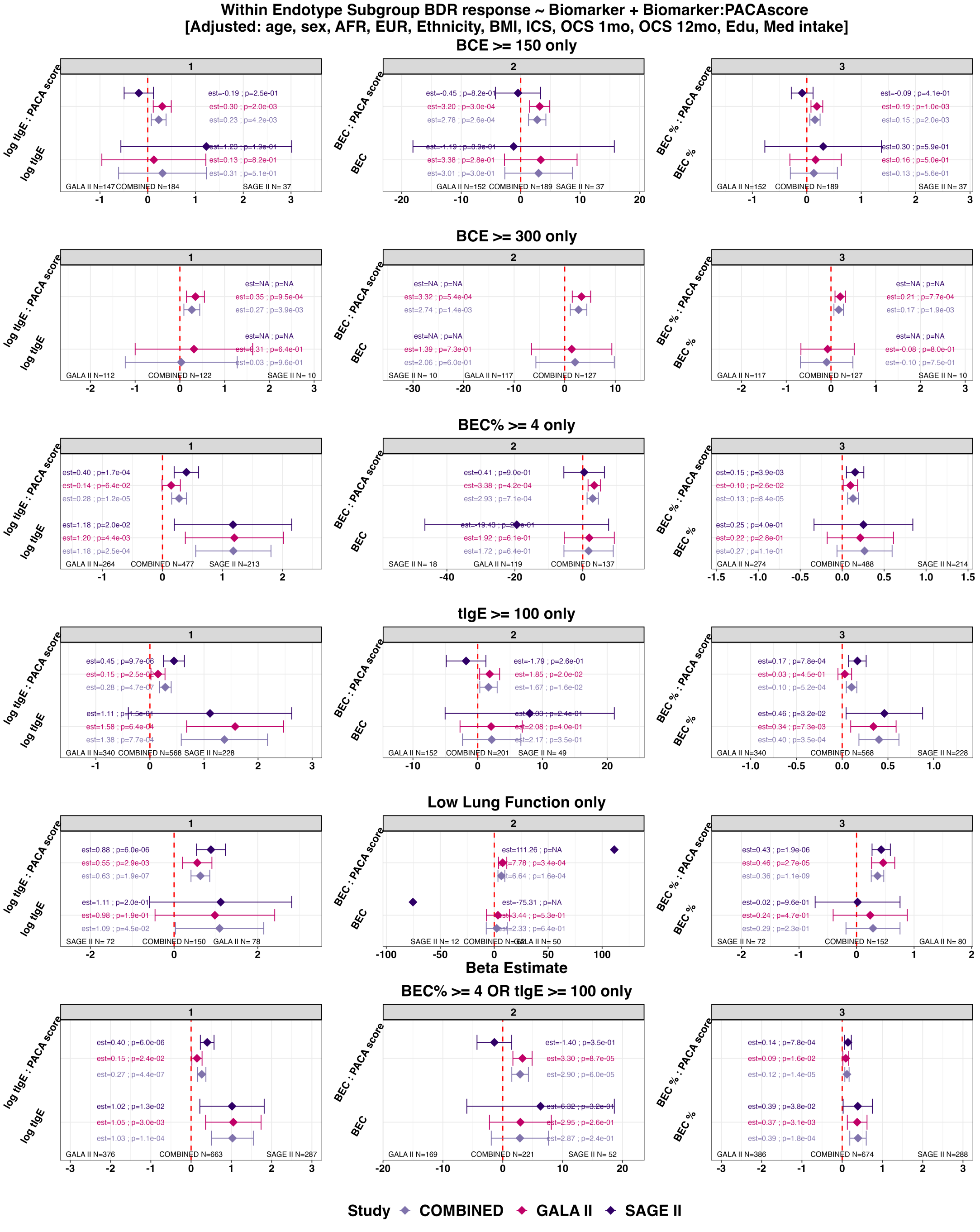

**Supplementary Figure S11:** Modeling BDR as a function of statistical interactions between the DNAm score and different biomarker definitions based on BEC and IgE, restricted to patient subgroups defined by different markers of T2-high status and baseline lung function. Each row presents results from linear regression models incorporating both a linear effect and an interaction term, using a different biomarker in each plot (BEC, BEC%, and IgE). Different rows display results evaluated in various patient subgroups, defined by different markers of T2-high status and baseline lung function. Presented as estimated effect sizes with 95% confidence intervals, results are stratified by study (GALA II, SAGE II) or combined (COMBINED. All models were adjusted for demographic and clinical covariates.

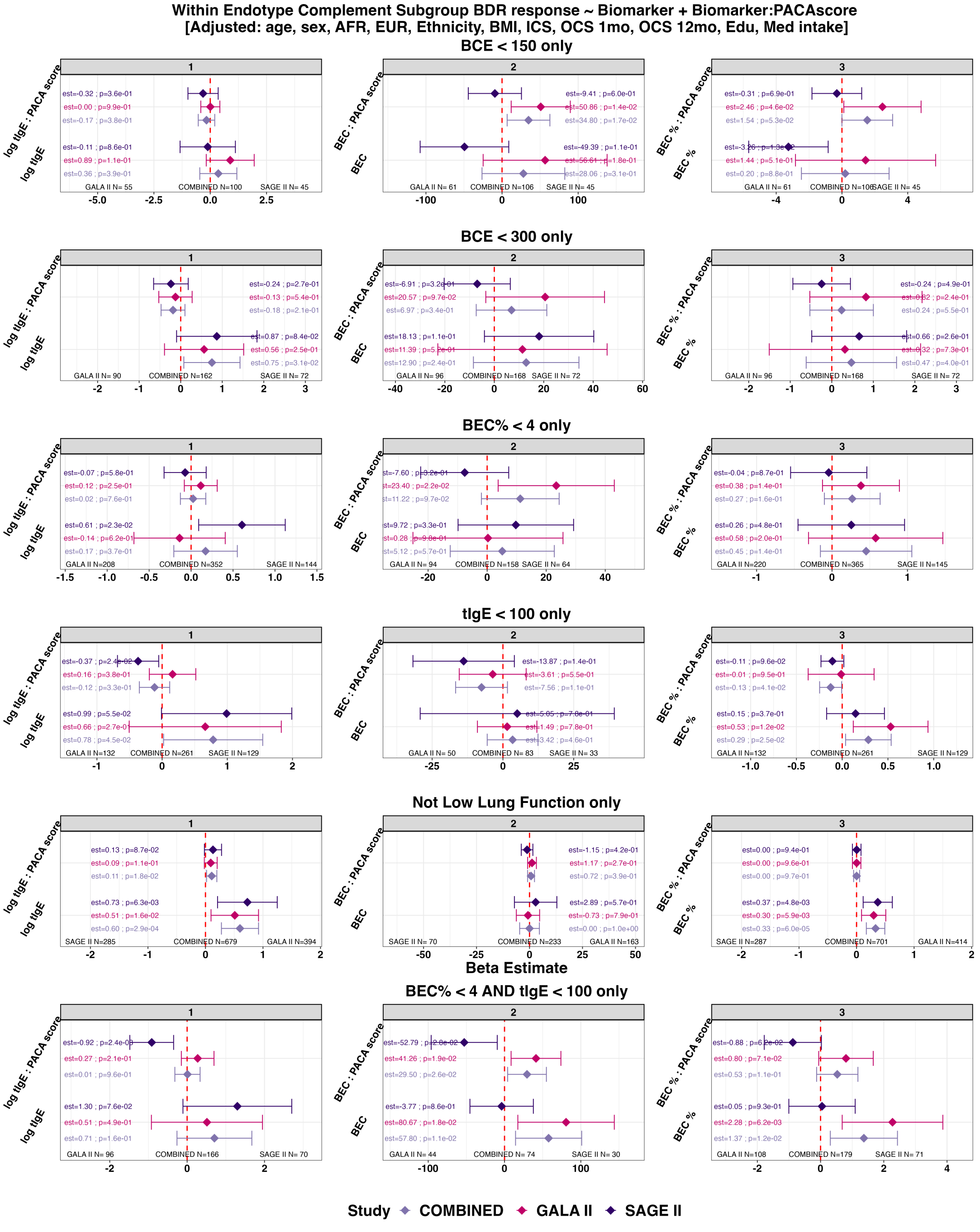

**Supplementary Figure S12:** Modeling BDR as a function of statistical interactions between the DNAm score and different biomarker definitions based on BEC and IgE, restricted to patient subgroups defined by different markers of T2-low status and baseline lung function. Each row presents results from linear regression models incorporating both a linear effect and an interaction term, using a different biomarker in each plot (BEC, BEC%, and IgE). Different rows display results evaluated in various patient subgroups, defined by different markers of T2-low status and baseline lung function. Presented as estimated effect sizes with 95% confidence intervals, results are stratified by study (GALA II, SAGE II) or combined (COMBINED. All models were adjusted for demographic and clinical covariates.

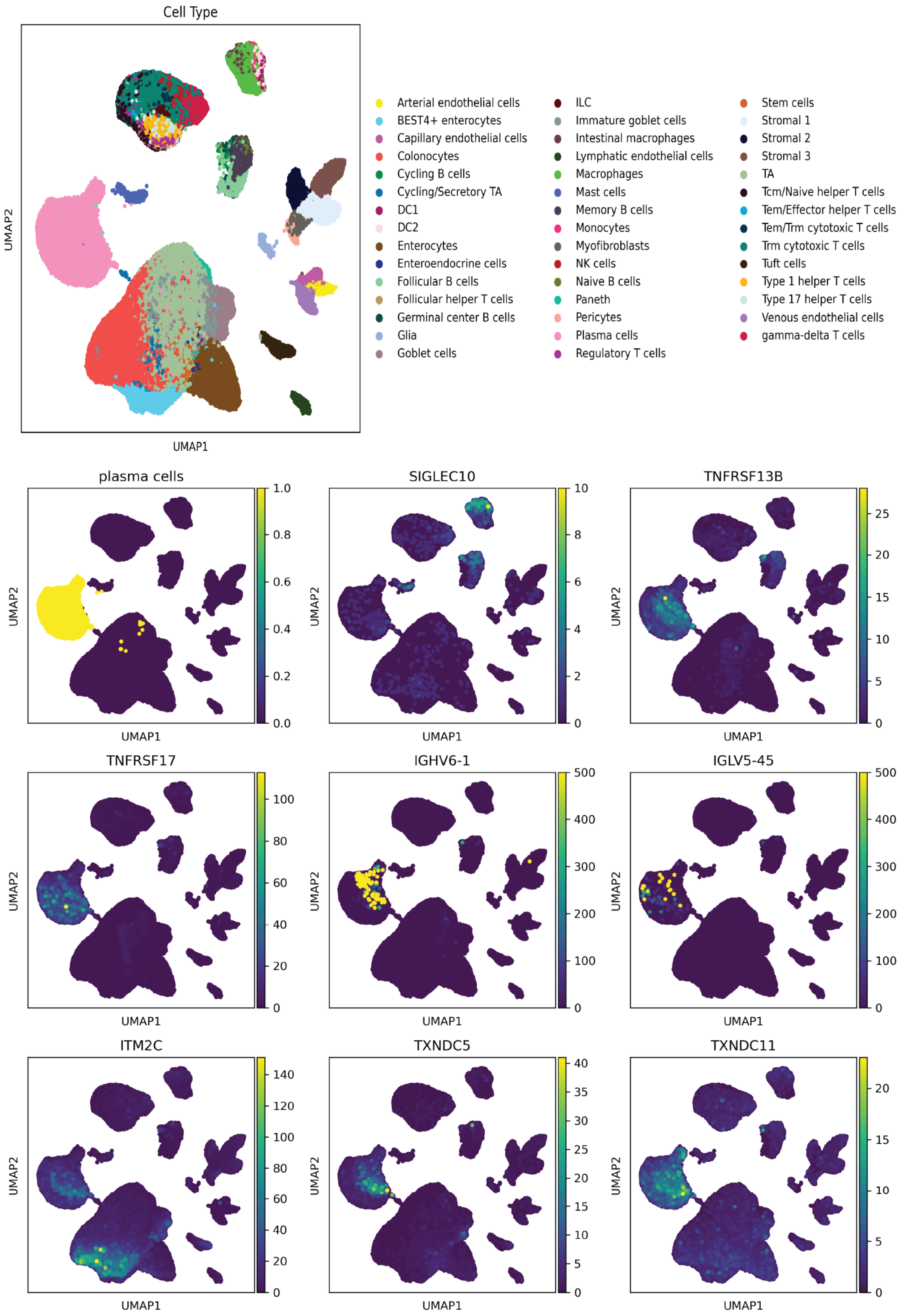

**Supplementary Figure S13:** UMAP embeddings of a single-cell model integrating four adult human intestine datasets, annotated by cell type and colored by expression of core T2ABC B cell-lineage genes.

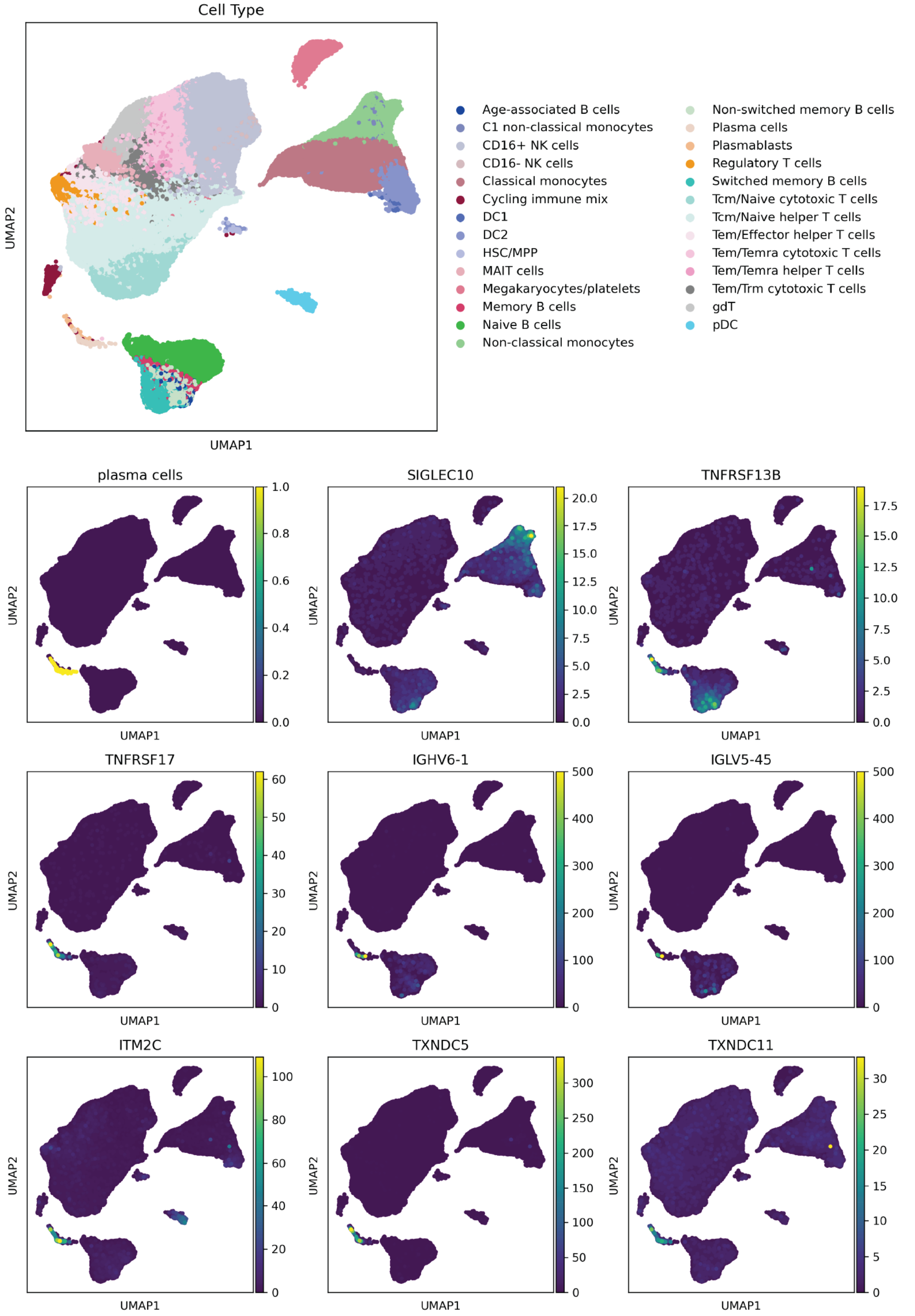

**Supplementary Figure S14:** UMAP embeddings of a single-cell model integrating four adult human blood datasets, annotated by cell type and colored by expression of core T2ABC B cell-lineage genes.

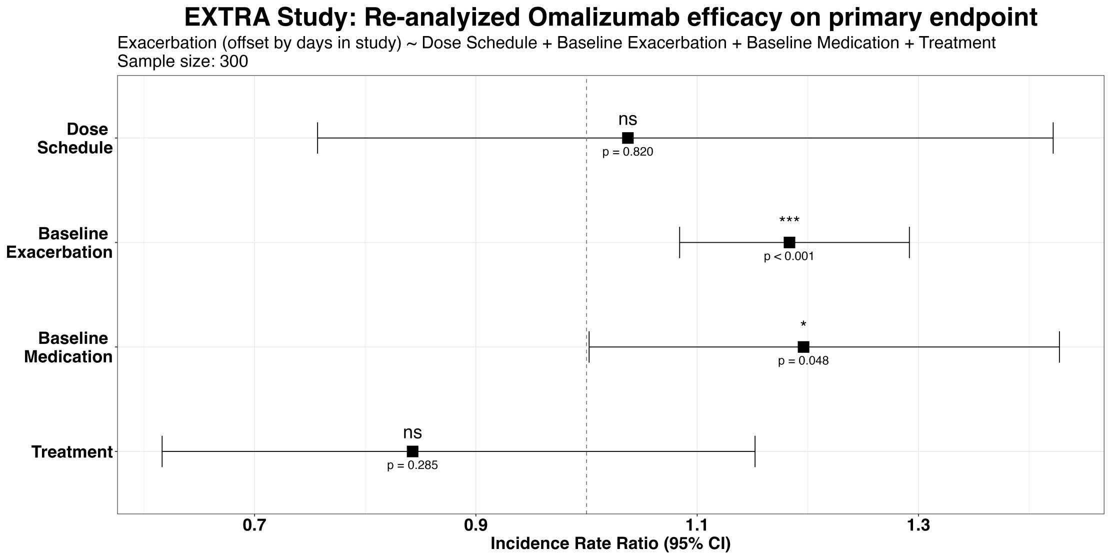

**Supplementary Figure S15:** Evaluation of the treatment effect of omalizumab on annualized exacerbation counts in patients with measured gene expression from the EXTRA randomized controlled trial. Incidence rate ratio (IRR) estimates, along with their corresponding 95% confidence intervals, are based on a Poisson regression model evaluating the treatment (placebo/omalizumab) effect on annualized exacerbation counts during the trial.

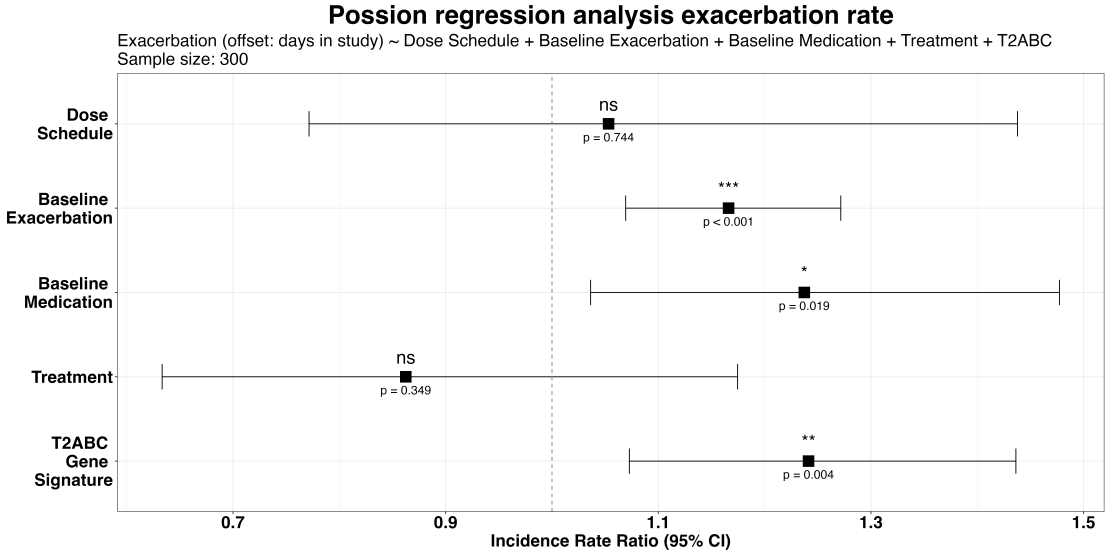

**Supplementary Figure S16:** Evaluation of the prognostic value of the T2ABC gene signature for predicting annualized exacerbation counts in patients with measured gene expression from the EXTRA randomized controlled trial. Incidence rate ratio (IRR) estimates, along with their corresponding 95% confidence intervals, are based on a Poisson regression model evaluating the effect of the T2ABC gene signature on annualized exacerbation counts during the trial.

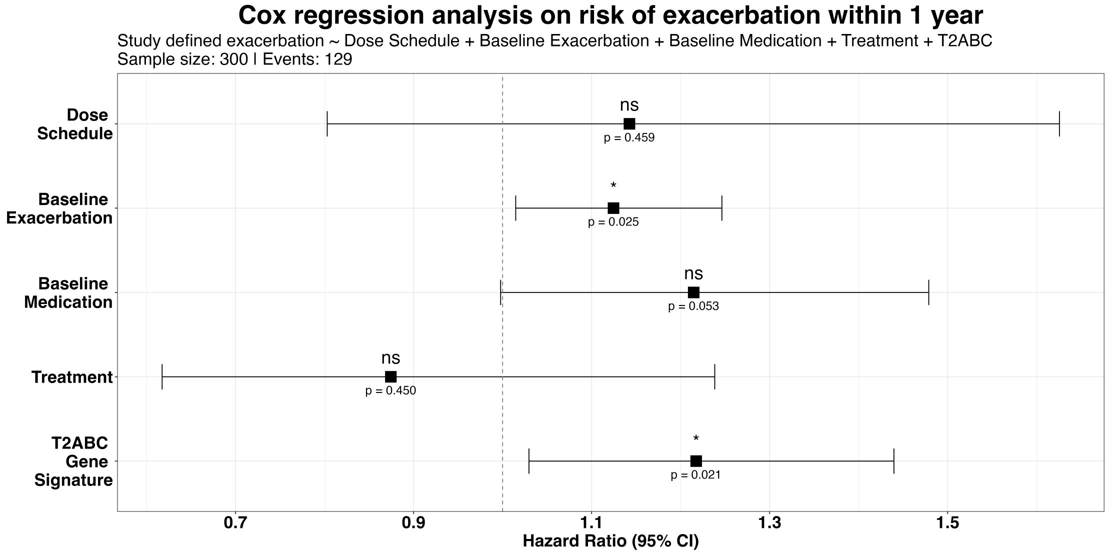

**Supplementary Figure S17:** Evaluation of the prognostic value of the T2ABC gene signature for predicting time to first exacerbation in patients with measured gene expression from the EXTRA randomized controlled trial. Hazard ratio (HR) estimates, along with their corresponding 95% confidence intervals, are based on a Cox regression model evaluating the effect of the T2ABC gene signature on time to first protocol-defined exacerbation during the trial.

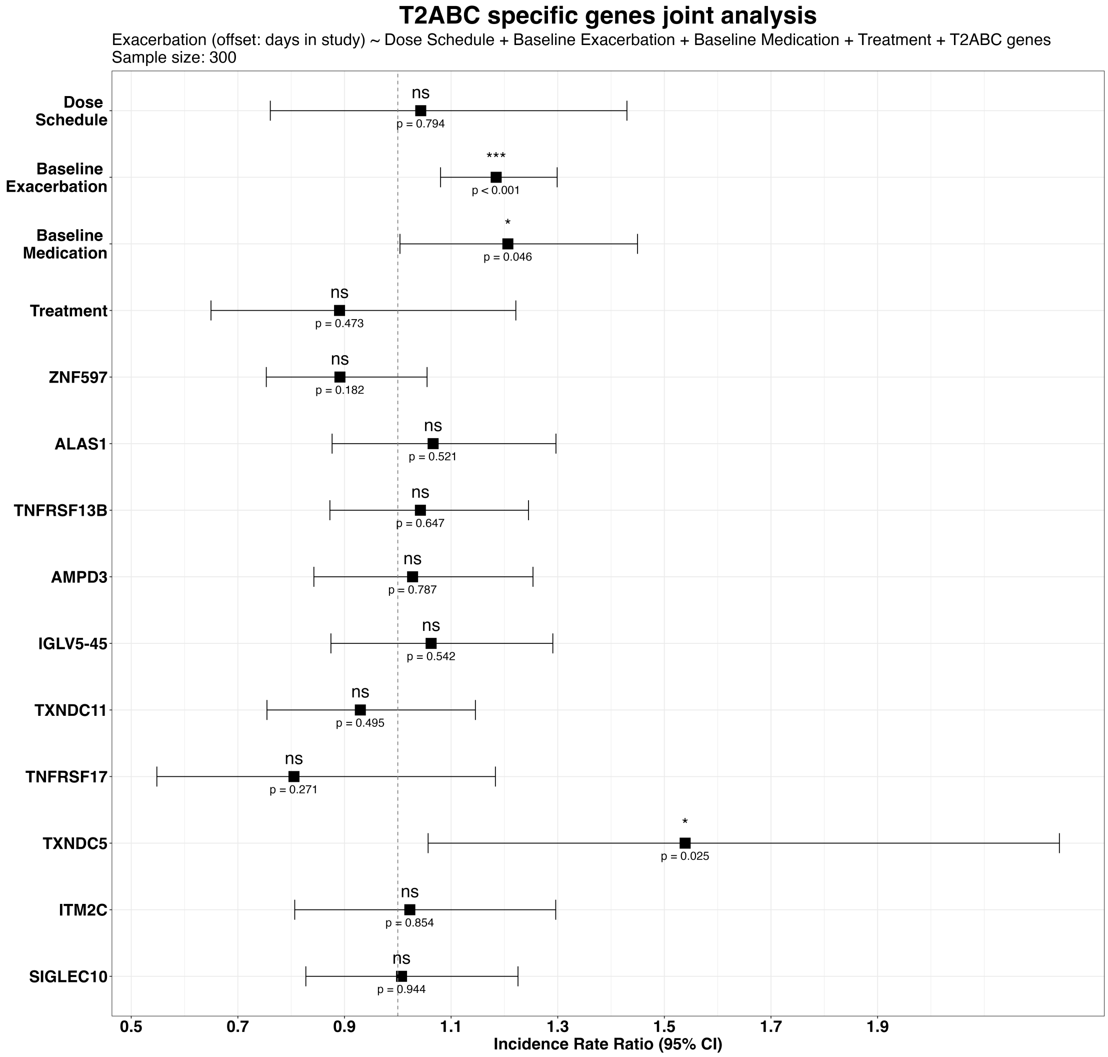

**Supplementary Figure S18:** A joint evaluation of the prognostic value of the ten genes composing the T2ABC expression score for predicting annualized exacerbation counts in patients with measured gene expression from the EXTRA randomized controlled trial. Incidence rate ratio (IRR) estimates, along with their corresponding 95% confidence intervals, are based on a Poisson regression model evaluating the effect of the ten genes on annualized exacerbation counts during the trial.

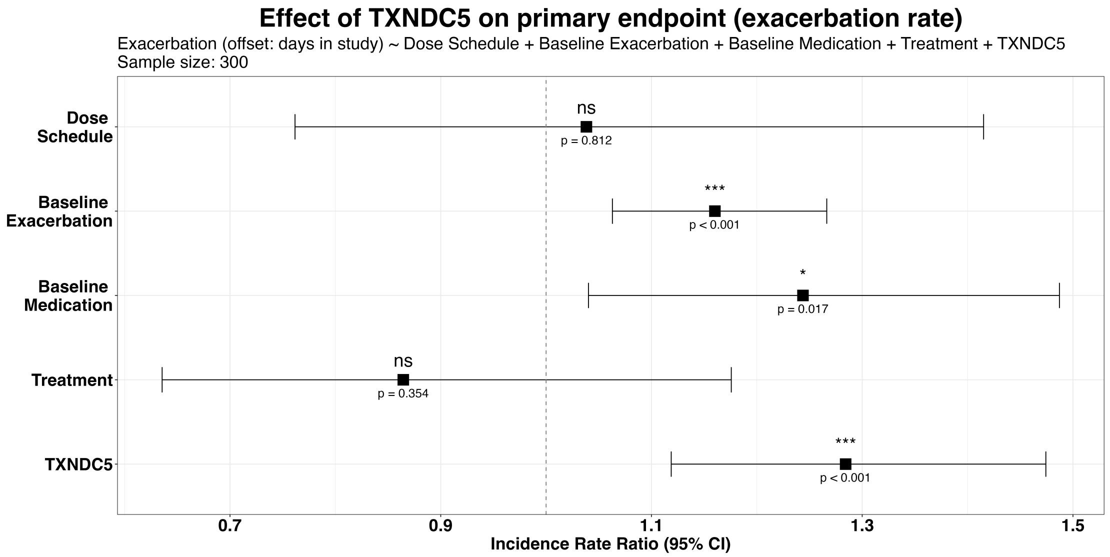

**Supplementary Figure S19:** Evaluation of the prognostic value of *TXNDC5* expression for predicting annualized exacerbation counts in patients with measured gene expression from the EXTRA randomized controlled trial. Incidence rate ratio (IRR) estimates, along with their corresponding 95% confidence intervals, are based on a Poisson regression model evaluating the effect of whole-blood *TXNDC5* expression on annualized exacerbation counts during the trial.

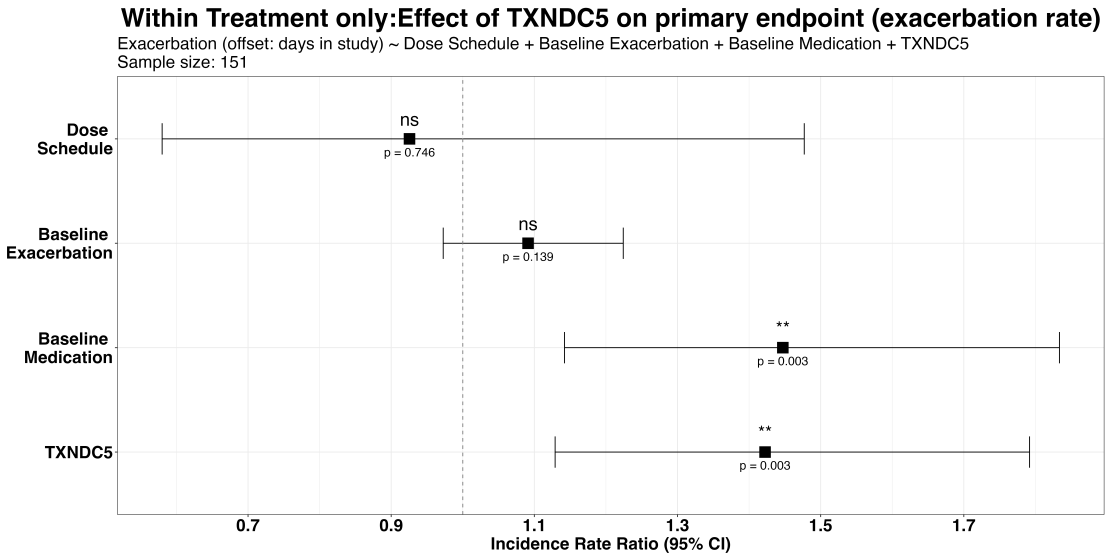

**Supplementary Figure S20:** Evaluation of the prognostic value of *TXNDC5* expression for predicting annualized exacerbation counts in omalizumab-treated patients with measured gene expression from the EXTRA randomized controlled trial. Incidence rate ratio (IRR) estimates, along with their corresponding 95% confidence intervals, are based on a Poisson regression model evaluating the effect of whole-blood *TXNDC5* expression on annualized exacerbation counts in omalizumab-treated patients during the trial.

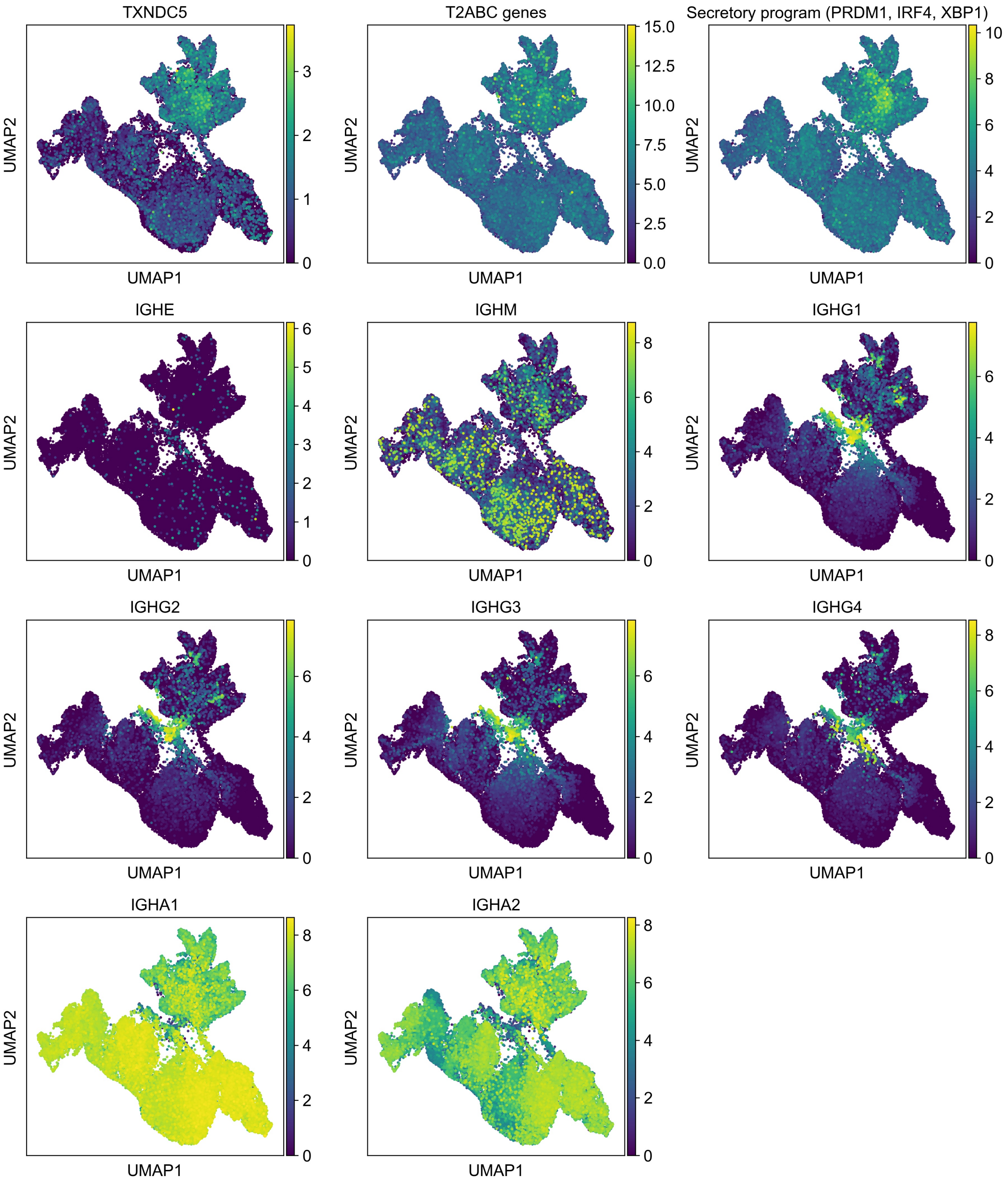

**Supplementary Figure S21:** UMAP embeddings of a single-cell model integrating plasma cells extracted from four adult human intestine datasets (a total of 46,348 cells), colored by expression of TXNDC5, the total expression across the seven predominantly plasma-cell expressed T2ABC genes, the total expression across canonical markers of the secretory program in plasma cells, and expression of immunoglobulin isotype-defining markers of plasma cells.

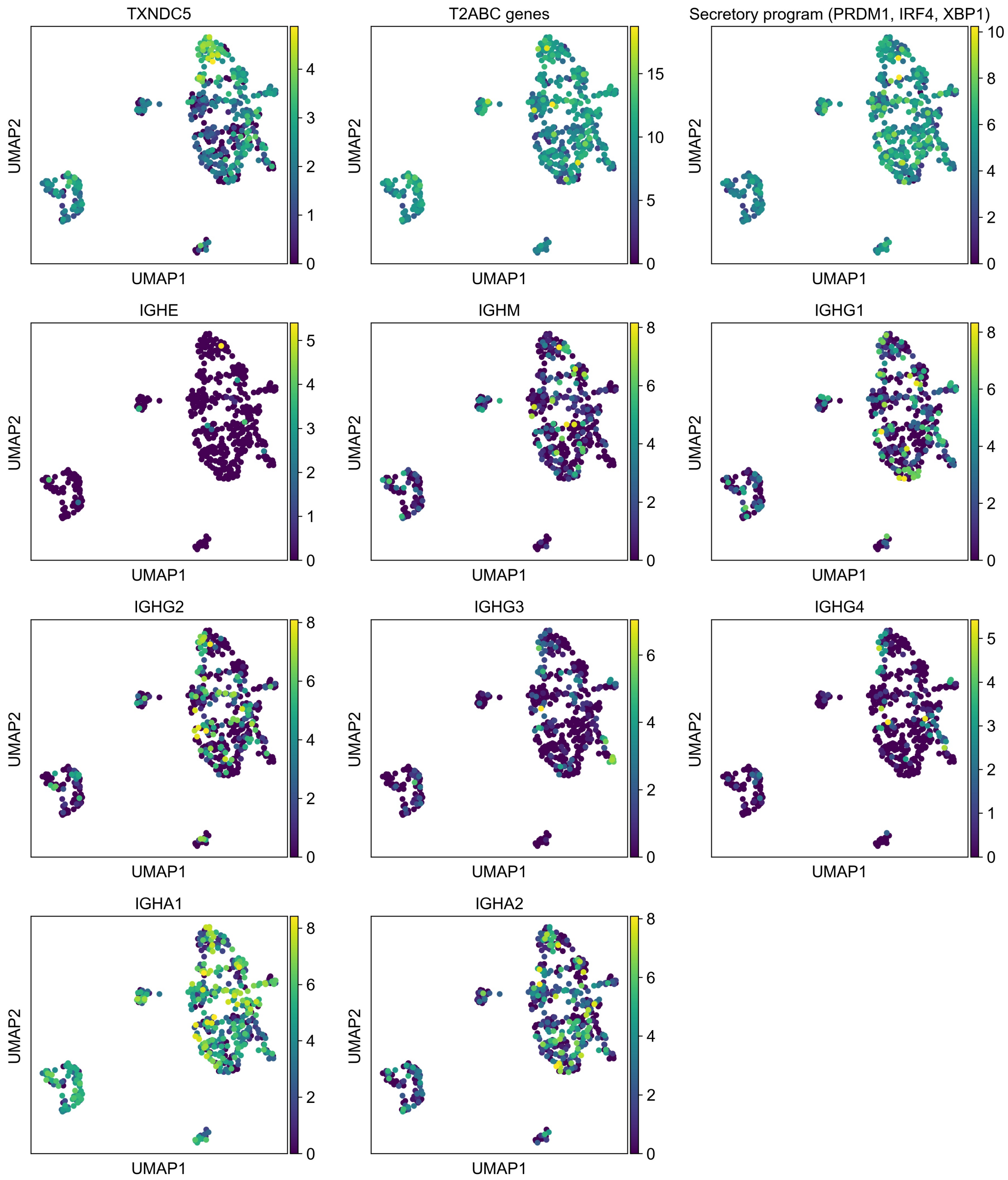

**Supplementary Figure S22:** UMAP embeddings of a single-cell model integrating plasma cells extracted from four adult human blood datasets (a total of 493 cells), colored by expression of TXNDC5, the total expression across the seven predominantly plasma-cell expressed T2ABC genes, the total expression across canonical markers of the secretory program in plasma cells, and expression of immunoglobulin isotype-defining markers of plasma cells.

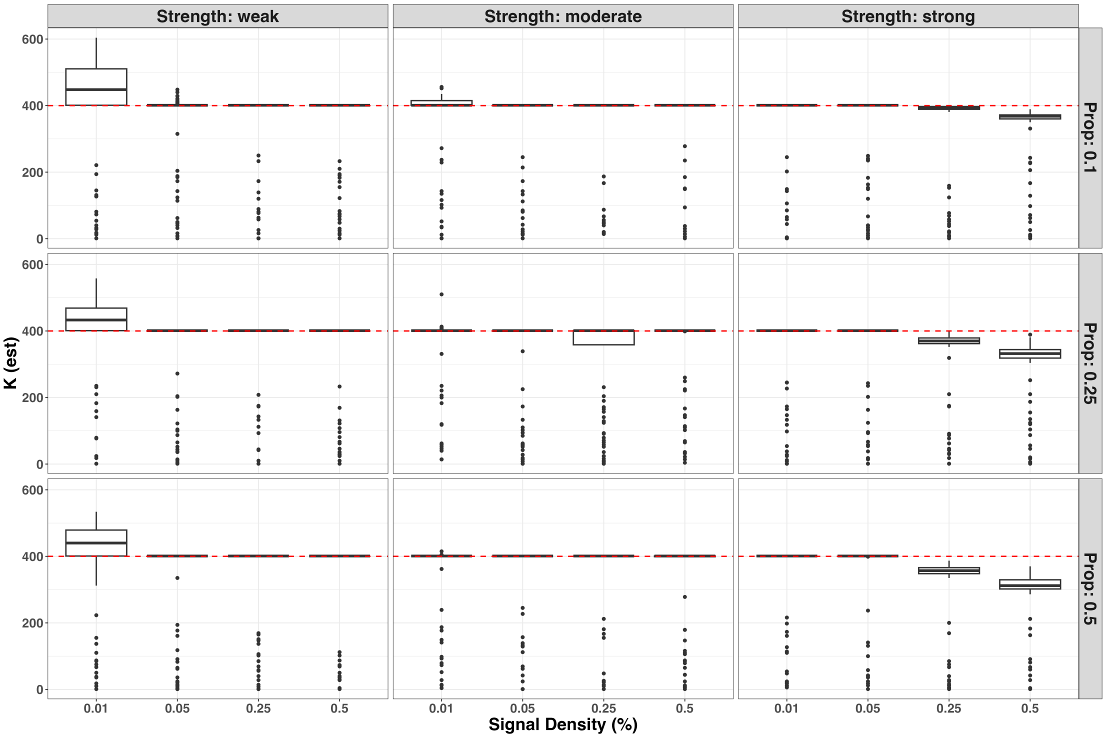

**Supplementary Figure S23:** Evaluation of the PACA-estimated dimension of shared variation ($k$) in synthetic data with two case subtypes. Results are based on simulated data with $n = 2000, k_0 = 400, m = 2000$; boxplots demonstrate the distribution of results across 100 simulations. Each column represents a different strength of the simulated subphenotypic signal, and each row represents a different proportion of case subtypes (Prop). The x-axis (signal density) reflects the number of features contributing to the subphenotypic signal (lower values indicate a more sparse signal).

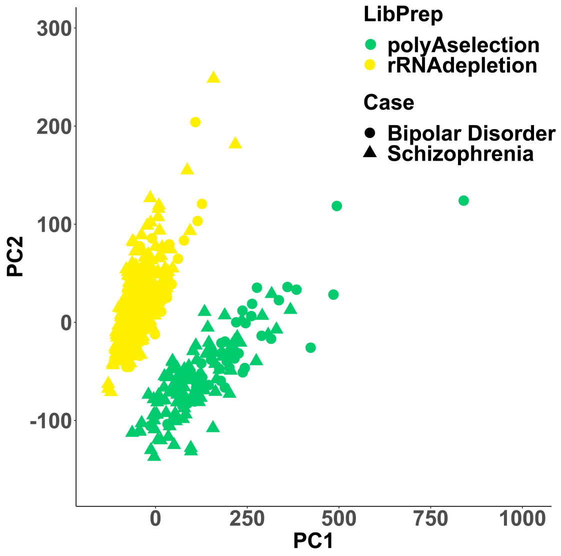

**Supplementary Figure S24:** The top two principal components (PCs) of a PCA applied to gene expression samples of schizophrenia (N=472) and bipolar disease (N=172) patients from the PsychENCODE study.

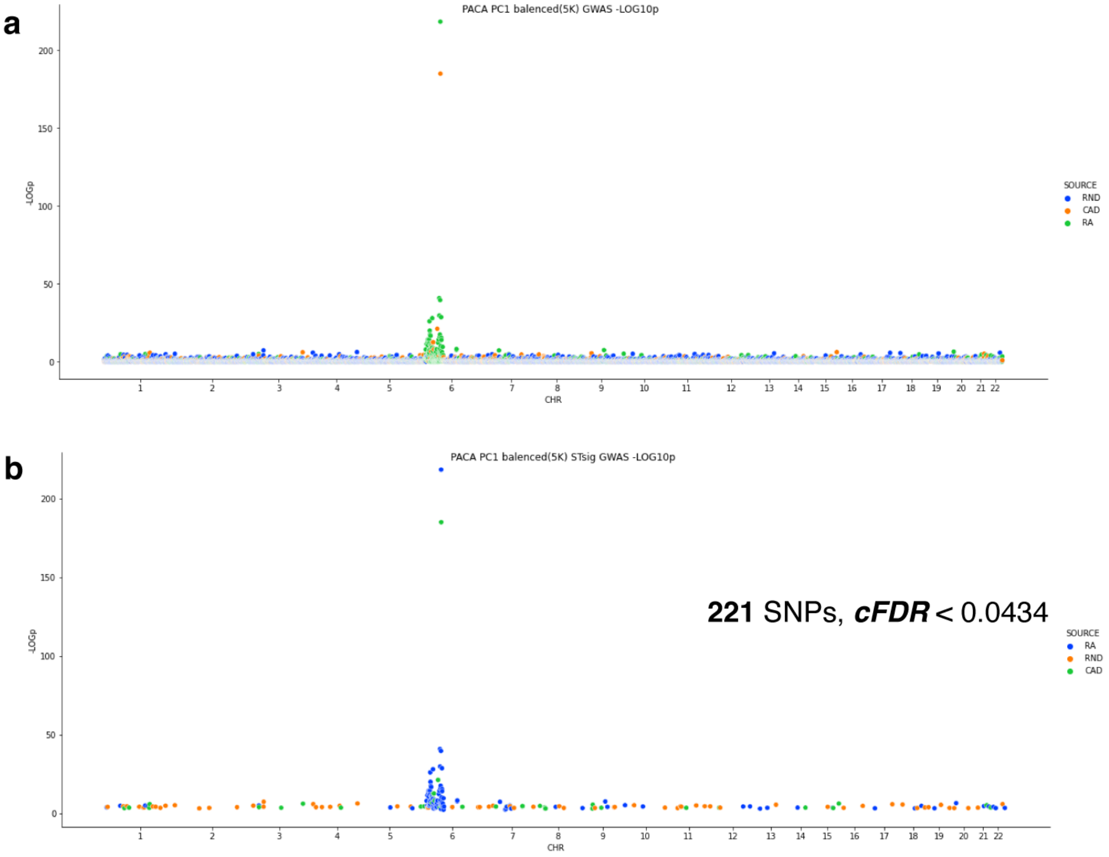

**Supplementary Figure S25:** Applying PACA to genotype data with a mixture of diseases from the UK BioBank. **(a)** Results of a GWAS with the top PACA component calculated from coronary artery disease (CAD) and rheumatoid arthritis (RA) patients (collectively treated as a single group of cases) and controls. The x-axis corresponds to chromosomal positions of the SNPs, and the y-axis shows their p-values on a -$\log_{10}$ scale. CAD (orange): significant SNPs from a CAD-specific GWAS; RA (green): significant SNPs from an RA-specific GWAS; RND (blue): random SNPs not associated with CAD or RA. **(b)** A visualization similar to (a), this time showing only the 221 SNPs passing Subtest cFDR (<0.034).
